# Impact of bariatric surgery on monthly earnings and employment: a national linked data study in England, 2014-2022

**DOI:** 10.1101/2025.02.05.25321712

**Authors:** Charlotte R. Bermingham, Daniel Ayoubkhani, Francesco Zaccardi, Karen Coulman, Jonathan Valabhji, Kamlesh Khunti, Dimitri J. Pournaras, Rita Santos, Nazrul Islam, Cameron Razieh, Ted Dolby, Vahé Nafilyan

**Affiliations:** Data and Analysis for Social Care and Health Division, Office for National Statistics, Newport, NP10 8XG, UK; Leicester Real World Evidence Unit, Diabetes Research Centre, University of Leicester, Leicester, LE5 4PW, UK; Population Health Sciences, Bristol Medical School, University of Bristol, Bristol, BS8 1NU; Department of Metabolism, Digestion and Reproduction, Faculty of Medicine, Chelsea and Westminster Hospital Campus, Imperial College London, London, SW10 9NH; NIHR Applied Research Collaboration. East Midlands, Diabetes Research Centre, University of Leicester, Leicester LE5 4PW; Department of Bariatric and Metabolic Surgery, North Bristol NHS Trust, Southmead Hospital, BS10 5NB, Bristol, UK; Centre for Health Economics, University of York, Alcuin Block A, YO10 5DD; Primary Care Research Centre, Faculty of Medicine, University of Southampton, Southampton SO16 5ST

## Abstract

**Objective:** Evaluate the impact of bariatric surgery on monthly earnings and employee status among working-age adults, and examine variations across sociodemographic characteristics.

**Design:** Retrospective longitudinal cohort study using national, linked administrative datasets.

**Setting:** Hospital inpatient services in England between 1 April 2014 and 31 December 2022.

**Participants:** 40,662 individuals who had a bariatric surgery procedure and obesity diagnosis during the study period, with no bariatric surgery history in the previous 5 years, and were 25 to 64 years old at the date of surgery. We also included 49,921 individuals sampled from the general population who had not had bariatric surgery matched by age and sex to those in the cohort who had bariatric surgery.

**Main outcome measures:** Monthly employee pay – for all months and only months where the individual was in paid employment – expressed in 2023 prices; paid employee status.

**Results:** Among people living with obesity who had bariatric surgery, there was a sustained increase in monthly employee pay from six months after surgery with a mean increase of £84 per month 5 years after surgery compared with the six months before surgery. Among those in paid employment, there was a sustained increase in the probability of being a paid employee from 4 months after bariatric surgery, with a mean increase of 4.3 percentage points 5 years after surgery. The increases in pay and probability of employment were greater for males. The increase in employee pay was not sustained over the 5-year follow up time for the youngest age groups.

**Conclusions:** Bariatric surgery is associated with an increased probability of being employed, resulting in increased earnings. These findings suggest that living with obesity negatively impacts labour market outcomes and that obesity management interventions are likely to generate economic benefits both to individuals and on a macroeconomic level by increasing the likelihood of employment of people living with obesity.

## Introduction

Poor health can affect an individual’s ability to work, impacting on employment, income, productivity, and overall wellbeing (1). There is some evidence that ill-health can affect labour market outcomes (2), but little evidence on the impact of health interventions in improving labour market outcomes of individuals. Demonstrating the effect of health interventions on economic outcomes could help increase the funding allocated to healthcare for the working age population and the labour market supply, as well as support economic growth.

Prevalence of obesity has been increasing globally (3). In England, 29% of adults aged 18 years and over were estimated to be living with obesity in 2022 and 64% living with overweight (including obesity) (4). Obesity is associated with a wide range of chronic conditions, including type 2 diabetes, sleep apnoea, heart failure and hypertension, as well as premature morbidity (5,6). The increase in risk of morbidity and mortality with increased BMI is larger among those under 50 years of age (6). Obesity can affect an individual’s labour market status through generally poorer health, via development of obesity related health conditions or through social discrimination (7). There is evidence that people living with obesity are less likely to be in paid employment (8), earn less on average than people not living with obesity, particularly among women (9), and have higher levels of sick leave (10,11) . Therefore, interventions to tackle obesity could improve labour market outcomes.

Bariatric surgery is the most effective obesity intervention for sustained weight-loss (12), leads to lower disease risk, particularly for type 2 diabetes, sleep apnea, nonalcoholic steatohepatitis and hypertension (13,14). There is also observational evidence of a beneficial effect across multiple outcomes such as cancer (15) and reduced all-cause mortality (16–18). It is therefore an ideal intervention to study the impact of weight-loss on employment. In the 2022-2023 financial year in England, ∼4,500 bariatric surgical procedures or gastric balloon procedures (not including revision procedures) were carried out (19). However, there is limited evidence on the impact of bariatric surgery and other obesity care interventions on labour market outcomes. There is significant interest from governments in understanding these impacts, for example in understanding the impacts of weight-loss drugs in the UK (20).

Previous studies have reported inconsistent findings on the impact of bariatric surgery on labour market outcomes; however, these have largely been based on small sample sizes with limited follow up (21–26). Nationwide studies using register data have been conducted in Scandinavian countries, where linked population-level employment and health datasets are established, and in Belgium using health insurance data. However, findings from these studies are inconclusive, with some reporting no changes in employment or earnings and others a positive or negative effect (27–30).

To our knowledge, no whole population study on the labour market impacts of bariatric surgery has been conducted in the UK. The UK has a publicly funded healthcare system that is free at the point of use. In addition, there are high levels of economic inactivity (people neither working nor seeking work) due to long-term ill health, which remains elevated compared with pre-pandemic levels (31). Therefore, this provides an ideal setting to investigate the labour market impacts of health interventions. Here, we used a population-level linked dataset for England, comprising electronic health records, sociodemographic information, and pay data for employees collected for tax purposes, to evaluate the long-term impact of bariatric surgery on pay and employment. We further examined heterogeneity in effects by sociodemographic characteristics.

## Methods

The aim of this study is to evaluate the effect of bariatric surgery on monthly earnings and employee status among working-age adults in England. We conducted a retrospective cohort study with a pre-post design: we analysed the change in monthly earnings and employee status following surgery, with individuals acting as their own controls, like in Self-Controlled Case Series (SCCS). We used a non-exposed group to control for time-varying confounders such as age and calendar time, as is best practice in SCCS (32).

### Study data

We used a linked, individual-level dataset for residents of England, combining: (1) Hospital Episode Statistics (HES) Admitted Patient Care (APC) records from 1 April 2009 to 31 December 2022 (33); (2) sociodemographic characteristics from Census 2011, or from Census 2021 if there was no link for the individual to Census 2011 (10% of individuals had no link to the 2011 Census) (34); (3) Office for National Statistics (ONS) death registrations for deaths that occurred from 1 January 2014 to 31 December 2022 and were registered by 31 December 2022. (35); (4) Office for National Statistics (ONS) birth registrations for births that occurred from 1 January 2014 to 31 December 2022 and were registered by 31 December 2023 (36); (5) Pay As You Earn (PAYE) Real Time Information (RTI) records from His Majesty’s Revenue and Customs (HMRC) covering 1 April 2014 to 31 December 2022. These are records of gross earnings paid to employees and recorded for tax purposes for the UK government, and are calendarised to monthly observations on employee status and pay (37).

The PAYE dataset was linked to the Census 2011 and 2021 through the ONS Demographic Index (38,39), which contains longitudinally linked administrative data to provide information on the population in England and Wales (40,41). The HES and death registration datasets were linked to the 2011 Census through the linkage of 2011 Census and NHS Patient Registers 2011-13 and the 2021 Census through the linkage of 2021 Census and 2021 NHS Patient Demographic Service (42). All datasets were then de-identified, prior to being harmonised and analysed.

### Study population and follow-up

We included individuals who had at least one episode in HES starting on or after 1 April 2014 and finishing on or before 31 December 2022, recording a primary or secondary Office of Population Censuses and Surveys (OPCS) code for bariatric surgery, concurrently with either a primary International Classification of Diseases (ICD-10) code for obesity or a secondary code for obesity and a primary code for an obesity-related condition (code lists in **Supplementary Table 1**). Individuals were excluded if they had at least one record in HES for a prior bariatric surgery procedure with episode end from 1 April 2009 and episode start on or before 31 March 2014. The procedure date of each individual’s first episode for bariatric surgery with an obesity diagnosis in the follow-up period was assigned as their index date.

An unexposed sample was created of individuals enumerated in the 2011 Census who did not have a record for bariatric surgery in HES either in the follow up time (episode end date from 1 April 2014 to 31 December 2022) or in the five years prior. The purpose of this sample was to adjust more accurately for time-varying confounders, which would be partially collinear with the within-individual treatment effect in the exposed cohort. In order to apply the age-based study inclusion criteria to the unexposed cohort, an index date was randomly assigned to individuals in the unexposed sample using the same distribution of operation dates as the bariatric surgery cohort (**Supplementary Text 1**). The unexposed population was sampled using stratified sampling by sex and five-year age band (for age at index date) to match the age-sex distribution in the bariatric surgery cohort.

We further restricted the sample to individuals who could be linked to a valid Census record for at least one of the 2011 or 2021 Censuses, were resident in England (based on the address recorded in HES on the index episode or, if not recorded, the address recorded in the linked Census data) and could be linked to at least one encrypted National Insurance number (NINo: the unique ID for each individual with data held by HMRC, including those paid via the PAYE system) and the Patient Registers 2011-13 (for individuals with a 2011 Census ID) or the 2019 Patient Demographic Service (for individuals with a 2021 Census ID) to enable linkage to the HES and deaths datasets. The sample was further restricted to individuals aged 25 to 64 years on the index date and who did not turn 65 of age before the end of that month, to include only individuals of working age and exclude the majority of students.

Individuals were followed up for a maximum of five years pre- and post-surgery, between 1 April 2014 and 31 December 2022 (the calendar time covered by reliable PAYE data for outcomes). Follow up time was right-censored at the earliest of death or turning age 69, and left-censored before turning age 21, to provide four years of potential follow up time before and after surgery.

The dataset linking and sample selection process for both exposed and unexposed cohorts is illustrated in **Supplementary Figure 1** and a sample flow is provided in **Supplementary Table 2.**

### Exposure and outcome variables

The exposure was time before or after the bariatric surgery procedure (exposure time), with the six months before surgery being the reference period. We included time periods up to five years before and after surgery, to enable testing for pre-surgery trends. Exposure time was included as monthly time periods for the month of surgery and the following five months to capture short-term effects after surgery; and as six-monthly periods thereafter to capture long-term effects. The time before surgery was also included in the exposure time variable as six-monthly periods. The month in which surgery occurred was defined as exposure month 1. The reference period, to which all time periods were compared, is the six months before surgery (exposure time 0). The exposure time was fixed at 0 for individuals in the unexposed sample.

Two outcomes were analysed: monthly employee pay (numerical: 2023 Sterling values); and paid employee status (numerical: 0/1). Monthly employee pay is gross pay recorded in PAYE. Monthly pay was winsorised at the 99.9% centile and deflated to 2023 prices using the Consumer Price Index including owner occupiers’ housing costs (CPIH). Being a paid employee was defined as receiving any monthly pay greater than zero. For the pay outcome, analyses were conducted on both the full dataset (employee pay overall) and on a dataset including only the months for which individuals were a paid employee (employee pay among those in employment).

### Statistical analysis

We described the data using either mean or percentage for a range of sociodemographic variables for the exposed and unexposed individuals, and compared the two groups using standardised mean differences. Standardised mean differences greater than 10% in absolute value indicated a large imbalance between the groups (43).

We compared the unadjusted outcomes (employee pay overall; employee pay among paid employees; paid employee status) over exposure time for the exposed and unexposed individuals. We also compared the unadjusted outcomes over calendar time (month-year) – including only the pre-surgery data for the exposed cohort to omit changes that may be due to the surgery – and by age at index date for each sex.

We analysed the change in monthly earnings and employee status following surgery, with individuals acting as their own controls. We fitted linear regression models (or linear probability models in the case of employment status as an outcome), with individual-level and calendar-time fixed effects; exposure time was included in the model with a reference level of the six-month period before surgery for the exposed group and all time periods for the unexposed group (**Supplementary Text 2**). Individual fixed effects capture confounding that does not vary within individuals over time; calendar month-year fixed effects were included as there could be factors which vary over calendar time for all individuals, such as background labour market conditions in the macroeconomy. The model was also adjusted for age in years (on the last day of each month) as a time-varying covariate, to account for changes due to ageing. Age was modelled as a natural cubic spline with four internal knots at the 20^th^, 40^th^, 60^th^ and 80^th^ centiles and boundary knots at the 10^th^ and 90^th^ centiles of its distribution; the number of knots was chosen by minimising the BIC.

We estimated robust standard errors accounting for within-individual correlation to accompany the estimated treatment effects, as well as 95% confidence intervals.

Information on all variables included in the models is reported in **Supplementary Table 3**. All analyses were carried out in R version 3.5.1. Dataset linkage and cleaning was carried out in SparklyR using Spark version 2.4.0. The statistical software package “lfe” version 3.0.0 was used to carry out the fixed effects linear regression modelling.

### Secondary analyses

We investigated heterogeneity of treatment effects across sociodemographic factors with an interaction between each sociodemographic variable of interest and the exposure. When investigating each sociodemographic factor as a potential effect modifier, we also included two-way interactions between the sociodemographic variable and each of age and calendar time to account for heterogenous time-varying confounding. The sociodemographic factors we investigated were: age at index date (in 10-year age bands, from 25-34 to 55-64 years), sex, region of residence in England, ethnic group, Index of Multiple Deprivation (IMD) quintile group and whether resident in a rural or urban area (**Supplementary Table 3**).

Giving birth can have a large effect on pay and employment (44), and the likelihood of birth may be different before and after surgery, we investigated the extent to which birth could affect the results by controlling for time-varying dummy variables (on a monthly basis) indicating whether an individual had given birth in the last year, or in the last five years not including the last year. As birth could also be hypothesised as a mediator rather than a confounder of the effect of bariatric surgery on pay and employment, in the main analysis it was not adjusted for.

### Sensitivity and placebo tests

We carried out sensitivity analyses to investigate the impact of changing the timing and duration of the reference time period, and to evaluate the extent to which changes in the outcomes due to the COVID-19 pandemic were accounted for (**Supplementary Text 3**).

We conducted placebo tests for which we would expect to observe null results (i.e., no trends in outcomes before or after surgery) if our models were working as intended, providing assurance that any non-null finding from the main analysis was a robust causal estimate (**Supplementary Text 4**).

### Patient and Public Involvement

Patients and the public were not involved in this research.

## Results

### Characteristics of the study population

We identified 43,968 individuals who had bariatric surgery with a hospital obesity diagnosis during follow-up, with no prior history of bariatric surgery. After limiting the cohort to those aged 25 to 64 years at the time of surgery, resident in England and who could link to census and PAYE datasets, 40,662 individuals (92.5%) remained for analysis (**Supplementary Table 2**). We included a cohort of 49,921 49,921 sex- and age-matched individuals in Census 2011 who had not had bariatric surgery (**Supplementary Table 3**).

The characteristics of the study population are shown in **Table 1** (variables are defined in **Supplementary Table 4**). Individuals who were living with obesity and had bariatric surgery had a mean age of 45 years, 80% were female and 85% were of White ethnicity. Compared with the cohort who had not had bariatric surgery, those who had a bariatric surgery procedure were broadly similar by ethnic group distribution, but were less likely of Asian and more likely of Black ethnicity. The largest differences between the two groups were observed for region of residence and deprivation level, with people who had bariatric surgery more likely to live in more deprived areas.

**Table 1:** Characteristics of the study population. Numbers are mean (standard deviation) or frequency (%).

| Characteristic | Level | Bariatric surgery | No bariatric surgery | Absolute standardised difference |
| --- | --- | --- | --- | --- |
| All people (n) | - | 40,662 | 49,921 |  |
| Age (mean, SD) |  | 45.4 (9.9) | 45.4 (10.0) | 0.1% |
| Sex (n, %) | Females | 32,344 (79.5) | 39,713 (79.6) | 0.0% |
| Ethnic group (n, %) | Asian | 1,650 (4.1) | 4,231 (8.5) | 23.9% |
|  | Black | 2,496 (6.1) | 1,874 (3.8) |  |
|  | Mixed | 1,002 (2.5) | 857 (1.7) |  |
|  | Other | 824 (2.0) | 461 (0.9) |  |
|  | White | 34,616 (85.1) | 42,252 (84.6) |  |
|  | Missing or not stated | 74 (0.2) | 246 (0.5) |  |
| Disability (n, %) | Yes - reduced a lot | 6,742 (16.6) | 2,397 (4.8) | 55.0% |
|  | Yes - reduced a little | 6,667 (16.4) | 3,165 (6.3) |  |
|  | No | 27,179 (66.8) | 44,113 (88.4) |  |
|  | Missing or not stated | 74 (0.2) | 246 (0.5) |  |
| IMD decile (n, %) | 1 (most deprived) | 5,552 (13.7) | 4,808 (9.6) | 36.0% |
|  | 2 | 6,122 (15.1) | 4,790 (9.6) |  |
|  | 3 | 5,767 (14.2) | 5,179 (10.4) |  |
|  | 4 | 4,885 (12.0) | 5,119 (10.3) |  |
|  | 5 | 4,089 (10.1) | 4,880 (9.8) |  |
|  | 6 | 3,749 (9.2) | 5,051 (10.1) |  |
|  | 7 | 3,240 (8.0) | 5,290 (10.6) |  |
|  | 8 | 2,807 (6.9) | 4,896 (9.8) |  |
|  | 9 | 2,547 (6.3) | 4,920 (9.9) |  |
|  | 10 (least deprived) | 1,904 (4.7) | 4,988 (10.0) |  |
| Region (n, %) | North East | 4,460 (11.0) | 2,308 (4.6) | 40.5% |
|  | North West | 2,881 (7.1) | 7,106 (14.2) |  |
|  | Yorkshire and The Humber | 2,834 (7.0) | 4,948 (9.9) |  |
|  | East Midlands | 1,980 (4.9) | 4,179 (8.4) |  |
|  | West Midlands | 4,988 (12.3) | 4,731 (9.5) |  |
|  | East of England | 3,281 (8.1) | 5,167 (10.4) |  |
|  | London | 8,623 (21.2) | 8,563 (17.2) |  |
|  | South East | 8,112 (19.9) | 8,181 (16.4) |  |
|  | South West | 3,503 (8.6) | 4,738 (9.5) |  |
| NS-SEC (n, %) | Higher managerial, administrative and professional occupations | 2,403 (5.9) | 5,245 (10.5) | 25.0% |
|  | Lower managerial, administrative and professional occupations | 8,637 (21.2) | 12,144 (24.3) |  |
|  | Intermediate occupations | 6,757 (16.6) | 8,305 (16.6) |  |
|  | Small employers and own account workers | 2,703 (6.6) | 3,575 (7.2) |  |
|  | Lower supervisory and technical occupations | 2,502 (6.2) | 2,593 (5.2) |  |
|  | Semi-routine occupations | 7,963 (19.6) | 7,530 (15.1) |  |
|  | Routine occupations | 4,423 (10.9) | 4,206 (8.4) |  |
|  | Never worked | 2,266 (5.6) | 2,146 (4.3) |  |
|  | Long-term unemployed | 1,086 (2.7) | 957 (1.9) |  |
|  | Full-Time students | 1,726 (4.2) | 2,785 (5.6) |  |
|  | Missing or not stated | 196 (0.5) | 435 (0.9) |  |
| Highest qualification<br>(n, %) | No qualifications | 6,423 (15.8) | 5,693 (11.4) | 25.5% |
|  | Level 1 qualifications | 7,449 (18.3) | 7,673 (15.4) |  |
|  | Level 2 qualifications | 8,353 (20.5) | 8,823 (17.7) |  |
|  | Apprenticeship | 682 (1.7) | 757 (1.5) |  |
|  | Level 3 qualifications | 6,272 (15.4) | 7,163 (14.3) |  |
|  | Level 4 qualifications and above | 9,691 (23.8) | 16,914 (33.9) |  |
|  | Other: Vocational/Work-related<br>qualifications/Foreign qualifications) | 1,596 (3.9) | 2,463 (4.9) |  |
|  | Missing or not stated | 196 (0.5) | 435 (0.9) |  |
| Rural/Urban (n, %) | Urban | 35,178 (86.5) | 42,233 (84.6) | 5.4% |

The overall median follow-up was 88 months, with a maximum of 105 months for both those who had bariatric surgery and those who did not. In those with bariatric surgery, median follow-up time was 48 months pre-surgery and 57 months post-surgery (**Supplementary Table 5**).

The most common bariatric procedures were Roux-en-Y bypass (20,427; 50.2%) and sleeve gastrectomy (15,329; 37.7%) (**Supplementary Table 6**). The number of procedures performed each month remained consistent until the COVID-19 pandemic, where large decreases were seen in the number of operations performed (**Supplementary Figure 2**).

### Unadjusted deflated employee pay and paid employee status

Among individuals who underwent bariatric surgery, the employment rate was 56.2% in the pre-surgery and 52.9% in the post-surgery period (**Supplementary Table 7**). Unadjusted median monthly deflated earnings (with no winsorisation applied) were £1,368 in the pre-surgery and £1,337 in the post-surgery period; corresponding figures when omitting months for which people were not in work were £2,437 and £2,528. Earnings and employment rate were higher among those who did not undergo bariatric surgery, with median monthly earnings of £1,828 overall and £2,901 among those in work only, and an overall employment rate of 64.7%. Employment rate and overall earnings were lower post-than pre-index date, as they were for those who underwent bariatric surgery.

The unadjusted monthly mean deflated employee pay for people who underwent bariatric surgery, overall and among those in work, followed similar trends in exposure time as those for people who did not undergo bariatric surgery. However, there was a reduction in the month of and following the operation for pay overall and among those in work, before a recovery to the pre-operation levels (**Figure 1**). There was a similar post-operation drop and recovery in the probability of employment for the people who underwent bariatric surgery. However, there was also a steeper decline in the probability of employment leading up to the operation than the overall decline in the probability of employment seen among the people who did not undergo bariatric surgery.

**Figure 1:**
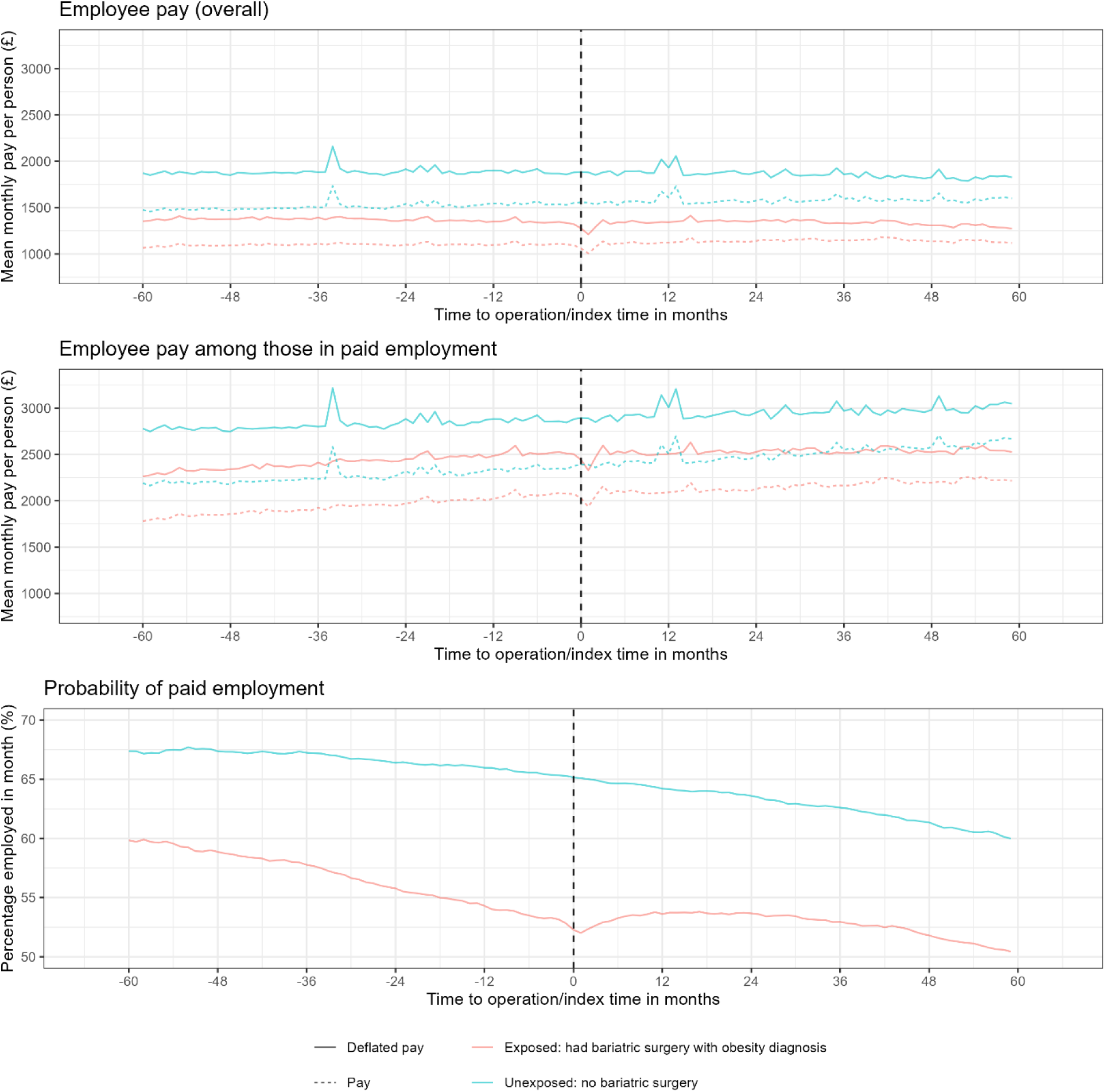
Unadjusted mean monthly employee pay and probability of being a paid employee over time before and after surgery for individuals with an obesity diagnosis who underwent bariatric surgery and individuals who did not undergo bariatric surgery 1. Pay is expressed in 2023 prices. 2. Month 0 is the month in which the surgery occurred for the treated cohort and a randomly assigned month for the unexposed sample.

Trends in pay by calendar time were broadly similar for people who underwent bariatric surgery (both overall and including only pre-operation time) and people who did not undergo bariatric surgery, but there were differences in trends of probability of employment by calendar time (**Supplementary Figure 3**; **Supplementary Text 5**). Trends of all outcomes by age, stratified by sex, also showed some differences (**Supplementary Figure 4; Supplementary Text 5**).

### Effect of bariatric surgery on employee pay and paid employee status

Bariatric surgery was associated with a sustained increase in the probability of being a paid employee (**Figure 2**; **Supplementary Table 8**). Compared to pre-surgery level, the probability of being a paid employee decreased by 0.8 (95% Cl: 0.6, 1.0) percentage points in the month after surgery, increasing to pre-surgery levels by the third month after the month of surgery. The employment probability then increased by 1.5 (95% Cl: 1.3, 1.8) percentage points in months 6-12 after surgery and continued increasing, reaching a maximum of 4.3 (3.7, 4.9) percentage points higher than pre-surgery levels in the months 54-60 after surgery. There was evidence of pre-surgery trends, with increased probabilities in the months before surgery compared to the 6 months immediately prior to surgery.

**Figure 2:**
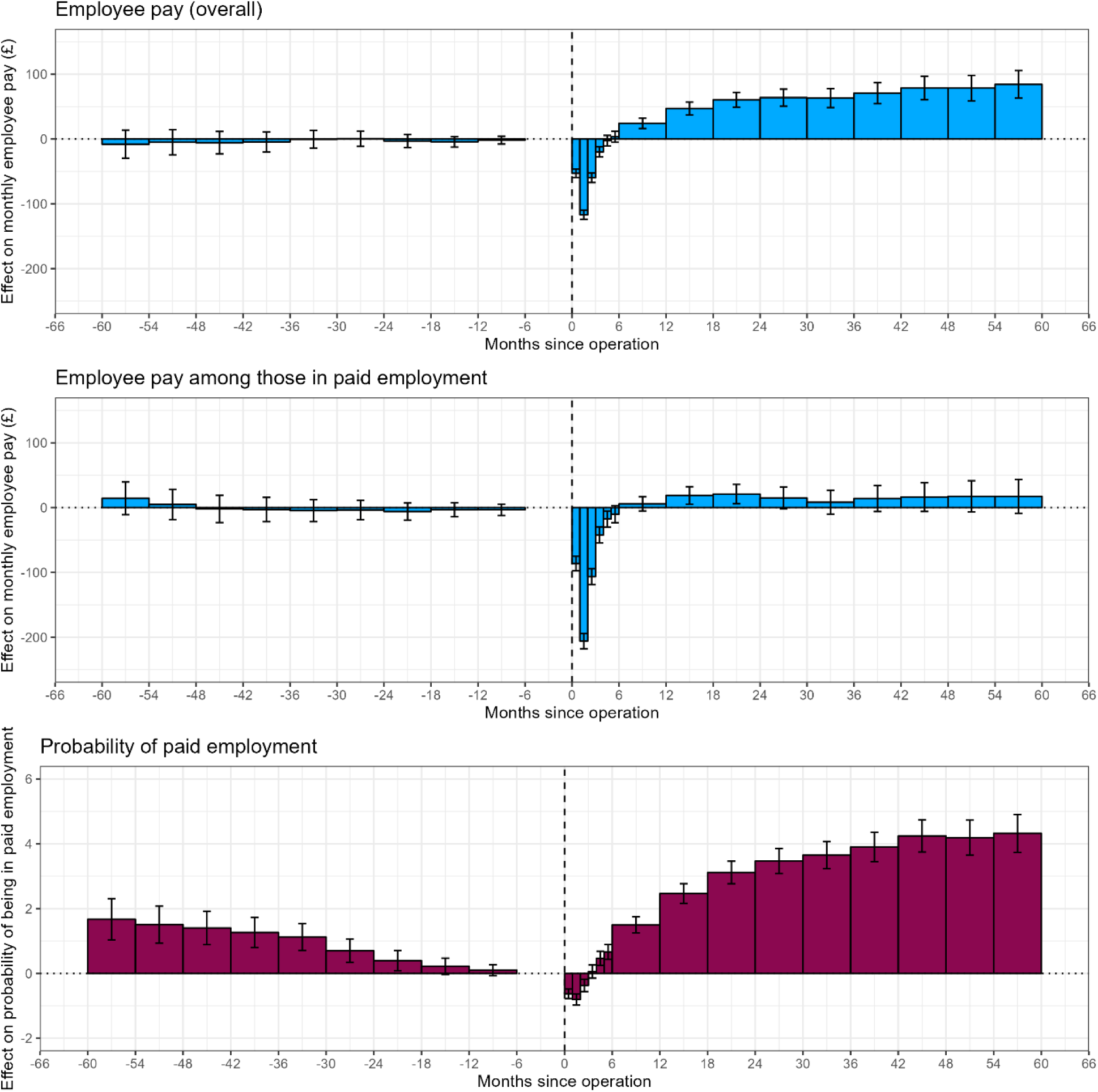
Effect of bariatric surgery on monthly employee pay and probability of being a paid employee before and after surgery 1. Month 0 to 1 is the month in which the surgery occurred for the treated cohort and a randomly assigned month for the unexposed sample. 2. Months -6 to 0 is the reference period to which all other time periods are compared.

Bariatric surgery was associated with a sustained average increase in employee pay, following a short decrease in pay in the first few months following surgery. Compared with the pre-surgery levels, employee earnings decreased in the month after surgery by an average of £117 (95% Cl: 110, 124), before increasing to pre-surgery levels by the fourth month after surgery (**Figure 2; Supplementary Table 9**). Compared to the six months before surgery, employee pay then increased to £61 (95% Cl: 49, 72) more than pre-surgery levels in months 18-24 after surgery, and continued to gradually increase compared to the six months before surgery to £84 (63, 106) more than pre-surgery levels in months 54-60 after surgery. There were no pre-surgery trends for employee pay overall.

By contrast, bariatric surgery was not strongly associated with an increase in pay among those in work (**Figure 2**; **Supplementary Table 10**). Pay among those in work initially decreased in the month after surgery, by an average of £206 (95% Cl: 194, 218) compared with pre-surgery levels, before increasing to pre-surgery levels by the fifth month after surgery. However, employee pay among those in work remained only slightly above or the same as pre-surgery levels in the following months, with a maximum of £21 (95% Cl: 6, 36) more than pre-surgery levels in months 18-24 after surgery.

There were no pre-surgery trends for employee pay overall and only a very small, possible downward trend in the pre-surgery trends for employee pay among those in work.

### Secondary analyses

For most of the treatment interactions investigated, the overall trends were broadly similar for each group, with an initial decrease followed by an increase in employee pay and probability of paid employment and no change in pay among those in paid employment.

However, there were differing trends by age group (**Supplementary Figures 5-7**). For the group of 25-34 years of age, there was an increase in pay from six-months after surgery, mirroring the main analysis, but this reduced over time, reaching similar and then slightly lower levels than pre-surgery. This trend was also seen in the group of 35-

44 years but at a later time after surgery. The probability of employment remained higher than pre-surgery levels for all age groups from six-months after surgery; however, for the group aged 25-34 years, pay among those in work decreased to lower than pre-surgery levels 24 months after surgery. Differences in pre-surgery trends may affect the results for this group.

The change in employee pay overall and employment after surgery was greater for men (**Supplementary Figures 8-10**). The probability of being a paid employee increased to a maximum of 5.8 percentage points higher than pre-surgery in months 54-60 in men compared to 3.9 in women. Employee pay for women plateaued at an average of £51 higher than pre-surgery from 18 months after surgery, whereas the effect for men was higher and continued increasing to reach £195 more than pre-surgery 54-60 months after surgery. Accounting for changes in birth rates among women before and after surgery had little impact on the overall estimates, with only a slight increase in pay overall, within the confidence limits of the main analysis (**Supplementary Table 11**).

Employee pay overall did not increase after bariatric surgery for people of Asian or Mixed ethnic groups and the probability of being a paid employee did not increase for people of Asian ethnicity (**Supplementary Figures 11-13**). However, the number of people in these ethnic groups is low, therefore the uncertainty in the results was higher.

The trends were broadly similar by area deprivation (IMD quintile), with a greater increase in the probability of paid employment from six-months after surgery for people living in the more deprived areas (**Supplementary Figures 14-16**). The trends were also broadly similar by region (**Supplementary Figures 17-19**). However, there were notable differences in some pre-surgery trends for the analyses by region and IMD, which may have influenced the results.

### Sensitivity analyses

Results from sensitivity analysis looking at the impact of the timing and length of the reference period were consistent with the findings of the main analysis (**Supplementary Text 3** and **Supplementary Tables 12-17**). There were slightly higher estimates of the association of surgery with the probability of being a paid employee when omitting data from the COVID-19 pandemic period, starting from 12 months after surgery (**Supplementary Table 18**). Results from placebo tests supported the robustness of the analysis method (**Supplementary Text 4** and **Supplementary Figures 20** and **21**).

## Discussion

### Main findings

In this nationwide study in England, in individuals who underwent a bariatric surgery procedure, we found an initial decrease in probability of employment immediately after surgery followed by a sustained increase compared with the six-month period before surgery. The increase in probability of employment was sustained from four months to five years after surgery (the end of the follow up period). The initial decrease followed by a sustained increase was also reflected in overall pay; however, there was little change in pay among only those in paid employment after the initial decrease, suggesting that the increase in pay was largely driven by the increased likelihood of being in paid employment, rather than changes in the rate of pay or hours worked.

Individuals were on average 3.7 percentage points more likely to be in paid employment 1-5 years after surgery, reaching a maximum of 4.3 percentage points more likely to be in paid employment 54 to 60 months after surgery. The average increase in monthly pay 1-5 years after surgery was £68, reaching £84 more than pre-surgery at 54 to 60 months after surgery. The total average cumulative increase in pay over 5 years after surgery was £3,180, which is a substantial part of the cost of surgery (45).

The size of the labour market effects of bariatric surgery differed between subgroups. In particular, the increase in pay and probability of employment after surgery was higher for men. However, trends were largely similar across the subgroups analysed, with the exception of age: the increase in pay seen from six months after surgery reduced to reach similar levels to pre-surgery after 30 and 48 months for the group of 25-34 and 35-44 years, respectively, and continued to decrease over time, whereas the increase in pay continued to rise over time for the 55-64 age group. In individuals between 25 and 34 years, a decline in pay among those in paid employment contributed to the decline in employee pay overall.

### Comparison with other studies

There is substantial evidence that a higher BMI is associated with lower pay, lower probability of employment, and high levels of sickness leave (8–11). However, evidence for the effect of bariatric surgery on these outcomes is more limited, often based on smaller sample sizes (21–26). A UK based study of 1,011 bariatric surgery patients found an increase in the number of patients in employment post-surgery (25). However, follow up was limited to 30 months and no comparison group who had not undergone bariatric surgery was included. To our knowledge, four nationwide, registry-based studies on the labour market effects of bariatric surgery have been carried out, with none in the UK. In Sweden, no change in pay or employment was observed over the 5 years after surgery (15,828 individuals who underwent bariatric surgery) (27); in Denmark, an increased probability of being in full-time employment 1-3 years after surgery was found but not in the longer term. There was a sustained increase in the probability of being in full-time employment for men, craftsmen and office workers (5,450 individuals who underwent bariatric surgery) (28). The third study, also conducted in Denmark, showed a lower risk of unemployment 5 years after surgery among men, but a higher risk in women and a higher risk of sickness absence up to 5 years after surgery in both men and women (10,328 individuals who underwent bariatric surgery) (30). A fourth study, in Belgium, found higher levels of employment 3 years after bariatric surgery (16,276 individuals who underwent bariatric surgery) (29).

Contrary to the Scandinavian registry-based studies, in our analysis we found a sustained increase in employment after bariatric surgery in both men and women; however, we did not observe a change in pay among those in employment. Some differences could be related to the national differences in healthcare or the labour market and welfare system.

### Possible mechanisms

The initial decrease in pay and employment is likely due to receiving sick leave pay that is lower than full pay. Statutory sick pay of £116.75 per week is paid for up to 28 weeks in the UK (46), therefore the lower likelihood of being in paid employment in the month of surgery and the following two months is likely related to people who have left the workforce or are in unstable employment, rather than people who are employed but are not receiving pay. The recovery time for starting to return to normal activities after bariatric surgery is 4 to 6 weeks (47), which is consistent with our results where the largest decrease in pay overall and among those in work is largest in the month after surgery, increasing to pre-surgery levels by six months after surgery.

The higher probability of being in paid employment after surgery, leading to increased overall pay, could be driven by an increase in general health due to weight loss, a lower likelihood of developing obesity related conditions, lower levels of social discrimination or stigma (48), or a combination of these factors, either enabling individuals who had left the workforce to return or decreasing the rate at which individuals are leaving the workforce.

At the same time, the decline in the probability of employment before bariatric surgery could be related to people being less likely to work while waiting for surgery due to poorer health (i.e., increasing disease severity while waiting for surgery). We do not account for this in our analyses because time-varying health status, which may account for these differences, could be a mediator of the impact of bariatric surgery on the outcomes. Instead, we showed that the results are sensitive to the timing of the baseline period, due to the pre-surgery trends, but the effect on post-surgery employment follows the same trends and was only slightly smaller than in our main results.

Reasons for differences in the increase in pay after bariatric surgery between men and women are not known. We found that, accounting for recent births, resulted only in a small change in the effect of bariatric surgery on the outcomes; therefore, factors other than differences in fertility after surgery are likely involved.

Some differences seen by ethnic group could be related to different eligibility criteria for bariatric surgery by ethnicity, with a lower BMI threshold determining procedure eligibility for people of South Asian, Chinese, other Asian, Middle Eastern, Black African or African-Caribbean family background (49).

It is possible that modelling trends in employment and pay is more challenging for younger people due to the volume of these people entering into the job market for the first time, therefore recording a large increase in pay, and higher differences between the unexposed and exposed samples, particularly in the 21-25 age range.

We found slightly higher increases in the probability of being a paid employee when omitting the COVID-19 pandemic period. This could be due to the pandemic having a greater impact on people living with obesity compared to the general population (50), resulting in differences in the trends of employment in calendar time. This is seen in the unadjusted trends of probability of employment in calendar time, with a larger reduction in employment rate for people who underwent bariatric surgery (before undergoing the surgery) compared with people who did not undergo bariatric surgery.

### Strengths and limitations of this study

To our knowledge, this is the largest population-wide study of labour market outcomes of bariatric surgery internationally, and the only such study outside of Scandinavia and Belgium. This investigation was made possible by a new national linked dataset comprising electronic health records, sociodemographic information and monthly pay records, which is the first linked dataset containing labour market and health data with near-complete coverage for individuals in England. The dataset also included a wide range of sociodemographic variables from the census to enable subgroup analysis.

We did not have reliable data on hours worked, therefore we could not distinguish changes in hours worked from changes in rate of pay. In addition, individuals who were self-employed were not included in the PAYE dataset and would be classed as not a paid employee in our analysis, along with people who were employed but not receiving pay (for example, due to maternity leave or sick leave). We also did not have information on benefits received, such as sick leave and disability benefit, which could be impacted by having bariatric surgery (26).

We used a sample of people from the general population who had not undergone bariatric surgery to better account for time-varying confounding, independently of the treatment effect (i.e. the effect of bariatric surgery). There were large differences between those who underwent bariatric surgery and those who did not for some sociodemographic characteristics, such as area-level deprivation, with a higher proportion of people who underwent bariatric surgery living in more deprived areas than in the general population. This may have contributed to the pre-surgery trends seen in some subgroups. An unexposed group constructed of the population eligible for bariatric surgery, but who did not undergo surgery, using information such as BMI (not available for this analysis) could be used as a comparison group, which may enable time-varying confounders (i.e. calendar time and ageing) to be better accounted for.

Bariatric surgery is highly effective, making it an ideal intervention to study the impact of weight-loss on employment outcomes. However, only a small percentage of the population eligible for bariatric surgery receive it, and the population who do differs from those eligible but who do not receive bariatric surgery (51). Further studies of alternative obesity care interventions, such as pharmaceutical interventions and weight-management programmes, are needed to enable comparisons between different interventions and employment outcomes.

## Conclusions

In our study, bariatric surgery resulted in a sustained increase in the probability of being a paid employee from four months after surgery over the five-year follow up period, leading to increased earnings. This suggests that living with obesity negatively impacts labour market outcomes and obesity care interventions are likely to generate substantial economic benefits by increasing earnings and employment of people living with obesity (52). The impact of bariatric surgery on an individual’s economic outcomes are not usually considered when calculating cost-effectiveness (53,54), or are estimated based on strong assumptions (11,24)(12,55). Estimates such as those produced by this analysis will aid in providing a more complete picture of the impact of bariatric surgery, supporting more robust health-economic appraisal (56).

## Data Availability

The source data used in this study is subject to controlled access due to its sensitive nature.

