## Supplementary Material for "Impact of bariatric surgery on monthly earnings and employment: a national linked data study in England, 2014-2022"

#### Supplementary Tables

**Supplementary Table 1:** Clinical codes used to define diagnoses and procedures in the Hospital Episode Statistics Admitted Patient Care dataset

| **Diagnosis or procedure** | **Codes** |
| --- | --- |
| **Bariatric surgery** | **OPCS-4 codes, primary and secondary procedures** |
| One-anastomosis gastric bypass | G281, G302, G304, G312 |
| Duodenal switch | G282, G283, G284, G716 |
| Sleeve gastrectomy | G285 |
| Gastric band | G303 |
| Roux-en-Y bypass | G321, G331 |
| Gastric balloon | G481, G485 |
| Endoscopic sleeve gastroplasty | G301 |
| **Primary diagnosis of obesity** | **ICD-10 codes, primary diagnosis**  E660, E661, E662, E668, E669 |
| **Secondary diagnosis of obesity** | **ICD-10 codes, secondary diagnosis**  E660, E661, E662, E668, E669 |
| **Primary diagnosis of an obesity related condition** | **ICD-10 codes, primary diagnosis** |
| Diaphragmatic hernia | K440, K441, K449 |
| Gastro oesophageal reflux | K210, K219 |
| Gastritis and duodenitis | K290-9 |
| Nausea vomiting | R11X |

**Supplementary Table 2:** Sample flow of individuals in the dataset who underwent bariatric surgery and had an obesity diagnosis

| **Sample selection criteria** | **Main sample (had bariatric surgery with an obesity diagnosis)** |
| --- | --- |
| **Is in HES dataset and has Census ID** (has at least one finished episode in HES APC where the episode started on or after 1 April 2014 and finished on or before 31 December 2022, and has a census ID for either 2011 or 2021 Census) | 43,238,218 |
| **Had bariatric surgery in follow up time** (has at least one episode where they had a metabolic surgery 1 April 2014 to 31 December 2022) | 57,787 |
| **No prior bariatric surgery** (does not have a record of a previous metabolic surgery with the episode ending on or after 1 April 2009 and starting on or before 31 March 2014) | 56,432 |
| **Has an obesity diagnosis** (has a primary diagnosis of obesity or a secondary diagnosis of obesity with a primary diagnosis of an obesity related condition) | 43,968 |
| **Resident in England** (as recorded in HES, or if not available then in the Census) | 43,701 |
| **Working age** (is aged 25 to 64 on the surgery date and does not turn 65 within the month of the surgery) | 40,754 |
| **Linkage to HMRC data** (has a valid encrypted National Insurance Number that can be linked to a census ID for either 2011 or 2021 Census) | 40,662 |

**Supplementary Table 3:** Sample flow of individuals in the dataset who did not undergo bariatric surgery

| **Sample selection criteria** | **Unexposed sample (no bariatric surgery)** |
| --- | --- |
| **Is enumerated in 2011 Census** | 56,963,120 |
| **No bariatric surgery in follow up time** (has not had any episode where they had metabolic surgery 1 April 2014 to 31 December 2022) | 56,910,963 |
| **No prior bariatric surgery** (does not have a record of a previous metabolic surgery (episode ending on or after 1 April 2009 and starting on or before 31 March 2014)) | 56,876,473 |
| **Can link to HES data** (has linkage to the Patient Register 2011-13) | 50,487,028 |
| **Resident in England** (as recorded in the Census) | 47,731,936 |
| **Working age** (is aged 25 to 64 on the index date and does not turn 65 within the month of the index date) | 25,353,079 |
| **Stratified sampling** (in the same proportions by sex and five-year age band as the sample of people who underwent bariatric surgery) | 50,001 |
| **Linkage to HMRC data** (has a valid encrypted National Insurance Number that can be linked to a census ID for either 2011 or 2021 Census) | 49,921 |

**Supplementary Table 4:** Variables used in the individual-month analysis dataset for modelling

| **Variable** | **Description** | **Source** |
| --- | --- | --- |
| **Outcomes** |  |  |
| Monthly employee pay (numerical) | Monthly employee pay is gross pay paid to employees as recorded in PAYE. Employees can be paid via the PAYE system according to a variety of payment frequencies: annual, quarterly, monthly, four-weekly, weekly, or irregularly. Payments were therefore calendarized such that the resulting linked dataset had a panel data structure, with monthly records for individuals. Monthly pay was imputed to be zero if it was negative. Where an individual had a Census ID that linked to multiple monthly PAYE records, the pay recorded in the PAYE RTI dataset was summed across all matching records for each month.  Monthly pay was wins~~z~~orised at the 99.9% centile and deflated to 2023 prices using the Consumer Price Index including owner occupiers' housing costs (CPIH). | PAYE RTI dataset |
| Employment (binary) | In employment was defined as receiving a monthly employee pay >£0. For the purposes of modelling, this was a numerical variable with value of either 0 or 1, rather than categorical, in order to produce probability estimates for the probability of being in paid employment. | PAYE RTI dataset |
| **Exposure** |  |  |
| Time since operation (categorical) | Time period since the month in which the bariatric surgery operation (see date of operation variable). The month of operation and the following 5 months are included as month time-periods, after which time periods are grouped into six monthly periods. Time periods before the month of operation are also grouped in six monthly periods, with time period 0 (the six months before the month of operation, set as the reference level for modelling.  For the unexposed dataset, the time since operation was set to zero (the reference level) throughout the dataset. | Hospital Episode Statistics (HES) |
| **Covariates** |  |  |
| Calendar time (categorical) | The month-year for all from April 2014 to December 2022 | - |
| Age (numerical) | Age in years on the last day of the month, calculated using Census date of birth | Census 2011 if available, Census 2021 if not |
| **Other variables** |  |  |
| Operation date (date) | The date of operation for the bariatric surgery procedure entry in the HES episode identified as being the first instance of a bariatric surgery procedure with an obesity diagnosis in HES for the individual in the exposed dataset (the index episode). Where an individual had multiple episodes that met the inclusion criteria, the episode with the earliest start date was used. Records were further deduplicated within individuals by taking the record with the earliest operation date and then the maximum episode key (the unique record identifier derived as part of HES processing) to obtain one episode per individual.  If no valid operation date was recorded for the relevant HES episode, the episode start date was used as the operation date, because the majority of bariatric surgery procedures occurred on the episode start date.  An operation date was assigned to individuals in the unexposed dataset for the purposes of calculating an age on index date for sample selection and to use for some descriptives and placebo tests. See **Supplementary Methods**. | HES |
| Age at operation (numerical) | The age in years on the date of operation, calculated using the Census date of birth and operation date variable. | Census 2011/2021, HES |
| Sex (binary) | Sex recorded in the Census (male/female) | Census 2011 if available, Census 2021 if not |
| 2011 LSOA of residence (categorical) | The 2011 LSOA recorded in the index episode for people in the exposed dataset. Where no LSOA was recorded, the LSOA from the linked Census entry was used. If the linked Census entry was from the 2021 Census, which records a 2021 LSOA, this was converted to the 2011 LSOA using a lookup from ONS Geography. | HES if available, Census 2011 if not, Census 2021 if not in either |
| Index of Multiple Deprivation decile (categorical) | The 2019 Index of Multiple Deprivation decile, linked to the 2011 LSOA of residence. | Census 2011 if available, Census 2021 if not, ONS Geography lookup |
| Index of Multiple Deprivation quintile (categorical) | The 2019 Index of Multiple Deprivation quintile, linked to the 2011 LSOA of residence. | Census 2011 if available, Census 2021 if not, ONS Geography lookup |
| Region (categorical) | The region of residence, linked to the 2011 LSOA of residence. | Census 2011 if available, Census 2021 if not, ONS Geography lookup |
| Rural/urban classification (binary) | The rural urban classification of the 2011 LSOA of residence. | Census 2011 if available, Census 2021 if not, ONS Geography lookup |
| Ethnic group (categorical) | Grouped ethnicity derived from the Census variable.  Values: White, Black, Asian, Mixed, Other, Missing or not stated. | Census 2011 if available, Census 2021 if not |
| Birth in last year (binary) | Time-varying flag to indicate whether the individual had given birth (live birth or stillbirth) in the last year. | ONS birth registrations |
| Birth in last 1 to 5 years (binary) | Time-varying flag to indicate whether the individual had given birth (live birth or stillbirth) between 1 and 5 years ago. | ONS birth registrations |

**Supplementary Table 5:** Follow up time, for the exposed individuals (had bariatric surgery with an obesity diagnosis) and the unexposed individuals (no bariatric surgery).

| **Dataset** | **Data** | **Minimum follow up (months)** | **Median follow up (months)** | **Maximum follow up (months)** |
| --- | --- | --- | --- | --- |
| Exposed only | Overall | 3 | 88 | 105 |
| Exposed only | Pre-surgery | 1 | 48 | 60 |
| Exposed only | Post-surgery | 1 | 57 | 60 |
| Unexposed only | Overall | 2 | 88 | 105 |

**Supplementary Table 6:** Total numbers of operations by type

| **Operation type** | **Count** | **Percentage** |
| --- | --- | --- |
| Roux-en-Y bypass | 20,426 | 50.2 |
| Sleeve gastrectomy | 15,329 | 37.7 |
| One-anastomosis gastric bypass | 1831 | 4.5 |
| Gastric band | 1429 | 3.5 |
| Gastric balloon | 1289 | 3.2 |
| Duodenal switch | 318 | 0.8 |
| Endoscopic sleeve gastroplasty | 40 | 0.1 |

**Supplementary Table 7:** Average pay ~~over whole datasets~~ before and after surgery, with no winsorisation applied, for the exposed individuals (had bariatric surgery with an obesity diagnosis) and the unexposed individuals (no bariatric surgery).

| **Individuals** | **Data** | **Median monthly deflated pay (overall) (£)** | **Median monthly deflated pay among those in work (£)** | **Maximum monthly deflated pay (£, nearest £100)** | **Percent in work (%)** |
| --- | --- | --- | --- | --- | --- |
| Exposed only | Overall | 1,351 | 2,485 | 2,722,400 | 54.4 |
| Exposed only | Pre-surgery | 1,368 | 2,437 | 2,471,700 | 56.2 |
| Exposed only | Post-surgery | 1,337 | 2,528 | 2,722,400 | 52.9 |
| Unexposed only | Overall | 1,878 | 2,901 | 8,628,700 | 64.7 |
| Unexposed only | Pre-index date | 1,889 | 2,839 | 8,628,700 | 66.5 |
| Unexposed only | Post-index date | 1,868 | 2,956 | 8,591,000 | 63.2 |

**Supplementary Table 8:** Model estimates for the probability of being in paid employment. Numbers in brackets indicate 95% CI.

| **Time since surgery (months)** | **Estimate (percentage points)** | **p value** |
| --- | --- | --- |
| -60 to -54 | 1.67 (1.03, 2.31) | <0.001 |
| -54 to -48 | 1.51 (0.94, 2.08) | <0.001 |
| -48 to -42 | 1.40 (0.89, 1.92) | <0.001 |
| -42 to -36 | 1.26 (0.80, 1.73) | <0.001 |
| -36 to -30 | 1.12 (0.71, 1.54) | <0.001 |
| -30 to -24 | 0.70 (0.34, 1.06) | <0.001 |
| -24 to -18 | 0.40 (0.08, 0.71) | 0.013 |
| -18 to -12 | 0.22 (-0.03, 0.47) | 0.085 |
| -12 to -6 | 0.10 (-0.07, 0.27) | 0.255 |
| 0 to 1 | -0.63 (-0.77, -0.48) | <0.001 |
| 1 to 2 | -0.80 (-0.97, -0.63) | <0.001 |
| 2 to 3 | -0.37 (-0.56, -0.18) | <0.001 |
| 3 to 4 | 0.06 (-0.15, 0.26) | 0.571 |
| 4 to 5 | 0.47 (0.25, 0.68) | <0.001 |
| 5 to 6 | 0.66 (0.43, 0.89) | <0.001 |
| 6 to 12 | 1.50 (1.25, 1.75) | <0.001 |
| 12 to 18 | 2.47 (2.16, 2.77) | <0.001 |
| 18 to 24 | 3.12 (2.77, 3.46) | <0.001 |
| 24 to 30 | 3.47 (3.08, 3.86) | <0.001 |
| 30 to 36 | 3.65 (3.23, 4.07) | <0.001 |
| 36 to 42 | 3.90 (3.45, 4.36) | <0.001 |
| 42 to 48 | 4.24 (3.75, 4.74) | <0.001 |
| 48 to 54 | 4.19 (3.65, 4.73) | <0.001 |
| 54 to 60 | 4.32 (3.74, 4.91) | <0.001 |

**Supplementary Table 9:** Model estimates for monthly employee pay. Numbers in brackets indicate 95% CI.

| **Time since surgery (months)** | **Estimate (£)** | **p value** |
| --- | --- | --- |
| -60 to -54 | -7.9 (-29.5, 13.8) | 0.475 |
| -54 to -48 | -4.9 (-24.4, 14.5) | 0.620 |
| -48 to -42 | -5.6 (-23.0, 11.7) | 0.526 |
| -42 to -36 | -4.5 (-19.9, 10.9) | 0.566 |
| -36 to -30 | -0.4 (-14.0, 13.3) | 0.959 |
| -30 to -24 | 0.5 (-11.2, 12.3) | 0.927 |
| -24 to -18 | -3.1 (-13.2, 7.0) | 0.550 |
| -18 to -12 | -4.3 (-12.4, 3.8) | 0.299 |
| -12 to -6 | -1.7 (-7.6, 4.3) | 0.579 |
| 0 to 1 | -52.9 (-59.4, -46.4) | <0.001 |
| 1 to 2 | -116.9 (-124.0, -109.8) | <0.001 |
| 2 to 3 | -59.7 (-67.2, -52.2) | <0.001 |
| 3 to 4 | -19.8 (-27.6, -12.0) | <0.001 |
| 4 to 5 | -2.4 (-10.5, 5.6) | 0.550 |
| 5 to 6 | 3.5 (-5.0, 12.0) | 0.420 |
| 6 to 12 | 24.2 (16.2, 32.2) | <0.001 |
| 12 to 18 | 47.1 (37.2, 57.0) | <0.001 |
| 18 to 24 | 60.5 (48.9, 72.1) | <0.001 |
| 24 to 30 | 63.9 (50.8, 77.1) | <0.001 |
| 30 to 36 | 63.2 (48.5, 77.8) | <0.001 |
| 36 to 42 | 70.8 (54.6, 87.0) | <0.001 |
| 42 to 48 | 78.7 (60.7, 96.7) | <0.001 |
| 48 to 54 | 78.6 (58.9, 98.3) | <0.001 |
| 54 to 60 | 84.4 (63.1, 105.8) | <0.001 |

**Supplementary Table 10:** Model estimates monthly employee pay among those in paid employment

| **Time since surgery (months)** | **Estimate (£)** | **p value** |
| --- | --- | --- |
| -60 to -54 | 14.4 (-10.8, 39.6) | 0.263 |
| -54 to -48 | 5.0 (-18.3, 28.3) | 0.676 |
| -48 to -42 | -2.0 (-23.0, 19.0) | 0.852 |
| -42 to -36 | -2.9 (-21.5, 15.8) | 0.762 |
| -36 to -30 | -4.6 (-21.5, 12.3) | 0.595 |
| -30 to -24 | -3.7 (-18.6, 11.2) | 0.627 |
| -24 to -18 | -6.0 (-19.3, 7.2) | 0.373 |
| -18 to -12 | -3.3 (-14.1, 7.5) | 0.544 |
| -12 to -6 | -3.3 (-12.1, 5.5) | 0.463 |
| 0 to 1 | -86.3 (-97.4, -75.1) | <0.001 |
| 1 to 2 | -206.0 (-217.8, -194.2) | <0.001 |
| 2 to 3 | -106.4 (-118.7, -94.2) | <0.001 |
| 3 to 4 | -42.1 (-54.3, -29.9) | <0.001 |
| 4 to 5 | -17.6 (-30.0, -5.1) | 0.006 |
| 5 to 6 | -10.1 (-23.2, 3.0) | 0.131 |
| 6 to 12 | 5.9 (-5.0, 16.8) | 0.289 |
| 12 to 18 | 18.8 (5.5, 32.2) | 0.006 |
| 18 to 24 | 21.0 (6.0, 35.9) | 0.006 |
| 24 to 30 | 14.9 (-2.0, 31.9) | 0.085 |
| 30 to 36 | 8.3 (-10.0, 26.7) | 0.373 |
| 36 to 42 | 14.1 (-5.9, 34.1) | 0.168 |
| 42 to 48 | 16.3 (-5.8, 38.5) | 0.149 |
| 48 to 54 | 17.5 (-6.6, 41.6) | 0.155 |
| 54 to 60 | 17.4 (-8.8, 43.5) | 0.194 |

**Supplementary Table 11:** Model estimate comparison table for the inclusion of birth variables (* = p value <0.05, ** = p value <0.01 and *** = p value < 0.001)

| **Time to/since surgery (months)** | **Main model pay** | **Main model pay with births in past year** | **Main model pay in work** | **Main model pay in work with births in past year** | **Main model employment** | **Main model employment with births in past year** |
| --- | --- | --- | --- | --- | --- | --- |
| -60 to -54 | -7.9  (-29.5, 13.8) | -6.2  (-27.8, 15.5) | 14.4  (-10.8, 39.6) | 16.5  (-8.7, 41.7) | 1.7***  (1.0, 2.3) | 1.7***  (1.1, 2.3) |
| -54 to -48 | -4.9  (-24.4, 14.5) | -3.6  (-23.0, 15.9) | 5.0  (-18.3, 28.3) | 7.0  (-16.3, 30.3) | 1.5***  (0.9, 2.1) | 1.5***  (1.0, 2.1) |
| -48 to -42 | -5.6  (-23.0, 11.7) | -4.2  (-21.6, 13.2) | -2.0  (-23.0, 19.0) | 0.3  (-20.6, 21.3) | 1.4***  (0.9, 1.9) | 1.4***  (0.9, 2.0) |
| -42 to -36 | -4.5  (-19.9, 10.9) | -3.0  (-18.4, 12.4) | -2.9  (-21.5, 15.8) | -0.7  (-19.3, 18.0) | 1.3***  (0.8, 1.7) | 1.3***  (0.8, 1.8) |
| -36 to -30 | -0.4  (-14.0, 13.3) | 1.2  (-12.5, 14.8) | -4.6  (-21.5, 12.3) | -2.2  (-19.1, 14.7) | 1.1***  (0.7, 1.5) | 1.2***  (0.7, 1.6) |
| -30 to -24 | 0.5  (-11.2, 12.3) | 1.9  (-9.8, 13.6) | -3.7  (-18.6, 11.2) | -2.0  (-16.9, 13.0) | 0.7***  (0.3, 1.1) | 0.7***  (0.4, 1.1) |
| -24 to -18 | -3.1  (-13.2, 7.0) | -2.1  (-12.2, 8.0) | -6.0  (-19.3, 7.2) | -4.6  (-17.9, 8.6) | 0.4*  (0.1, 0.7) | 0.4**  (0.1, 0.7) |
| -18 to -12 | -4.3  (-12.4, 3.8) | -3.6  (-11.7, 4.5) | -3.3  (-14.1, 7.5) | -2.1  (-12.9, 8.7) | 0.2  (-0.0, 0.5) | 0.2  (-0.0, 0.5) |
| -12 to -6 | -1.7  (-7.6, 4.3) | -1.4  (-7.3, 4.6) | -3.3  (-12.1, 5.5) | -2.7  (-11.6, 6.1) | 0.1  (-0.1, 0.3) | 0.1  (-0.1, 0.3) |
| 0 to 1 | -52.9***  (-59.4, -46.4) | -53.1***  (-59.6, -46.5) | -86.3***  (-97.4, -75.1) | -86.5***  (-97.6, -75.4) | -0.6***  (-0.8, -0.5) | -0.6***  (-0.8, -0.5) |
| 1 to 2 | -116.9***  (-124.0, -109.8) | -117.1***  (-124.2, -110.0) | -206.0***  (-217.8, -194.2) | -206.4***  (-218.2, -194.6) | -0.8***  (-1.0, -0.6) | -0.8***  (-1.0, -0.6) |
| 2 to 3 | -59.7***  (-67.2, -52.2) | -59.9***  (-67.4, -52.4) | -106.4***  (-118.7, -94.2) | -106.8***  (-119.1, -94.6) | -0.4***  (-0.6, -0.2) | -0.4***  (-0.6, -0.2) |
| 3 to 4 | -19.8***  (-27.6, -12.0) | -20.1***  (-27.8, -12.3) | -42.1***  (-54.3, -29.9) | -42.6***  (-54.8, -30.4) | 0.1  (-0.1, 0.3) | 0.0  (-0.2, 0.3) |
| 4 to 5 | -2.4  (-10.5, 5.6) | -2.8  (-10.8, 5.3) | -17.6**  (-30.0, -5.1) | -18.1**  (-30.5, -5.6) | 0.5***  (0.2, 0.7) | 0.5***  (0.2, 0.7) |
| 5 to 6 | 3.5  (-5.0, 12.0) | 3.2  (-5.3, 11.7) | -10.1  (-23.2, 3.0) | -10.6  (-23.8, 2.5) | 0.7***  (0.4, 0.9) | 0.6***  (0.4, 0.9) |
| 6 to 12 | 24.2***  (16.2, 32.2) | 23.8***  (15.8, 31.8) | 5.9  (-5.0, 16.8) | 5.2  (-5.7, 16.1) | 1.5***  (1.3, 1.7) | 1.5***  (1.2, 1.7) |
| 12 to 18 | 47.1***  (37.2, 57.0) | 46.9***  (37.0, 56.8) | 18.8**  (5.5, 32.2) | 18.6**  (5.2, 31.9) | 2.5***  (2.2, 2.8) | 2.4***  (2.1, 2.8) |
| 18 to 24 | 60.5***  (48.9, 72.1) | 60.9***  (49.3, 72.5) | 21.0**  (6.0, 35.9) | 22.1**  (7.2, 37.0) | 3.1***  (2.8, 3.5) | 3.1***  (2.8, 3.5) |
| 24 to 30 | 63.9***  (50.8, 77.1) | 64.6***  (51.5, 77.7) | 14.9  (-2.0, 31.9) | 17.0*  (0.1, 34.0) | 3.5***  (3.1, 3.9) | 3.5***  (3.1, 3.8) |
| 30 to 36 | 63.2***  (48.5, 77.8) | 63.9***  (49.3, 78.5) | 8.3  (-10.0, 26.7) | 10.3  (-8.0, 28.6) | 3.7***  (3.2, 4.1) | 3.6***  (3.2, 4.1) |
| 36 to 42 | 70.8***  (54.6, 87.0) | 71.5***  (55.4, 87.7) | 14.1  (-5.9, 34.1) | 15.6  (-4.4, 35.6) | 3.9***  (3.4, 4.4) | 3.9***  (3.4, 4.4) |
| 42 to 48 | 78.7***  (60.7, 96.7) | 79.5***  (61.5, 97.5) | 16.3  (-5.8, 38.5) | 18.1  (-4.0, 40.2) | 4.2***  (3.7, 4.7) | 4.2***  (3.8, 4.7) |
| 48 to 54 | 78.6***  (58.9, 98.3) | 79.7***  (60.0, 99.4) | 17.5  (-6.6, 41.6) | 19.9  (-4.2, 44.0) | 4.2***  (3.7, 4.7) | 4.2***  (3.7, 4.7) |
| 54 to 60 | 84.4***  (63.1, 105.8) | 85.5***  (64.2, 106.8) | 17.4  (-8.8, 43.5) | 19.5  (-6.6, 45.7) | 4.3***  (3.7, 4.9) | 4.3***  (3.8, 4.9) |
| Birth in last year |  | -319.7***  (-376.8, -262.5) |  | -722.6***  (-811.4, -633.8) |  | -6.2***  (-8.0, -4.4) |
| Birth in last 1-5 years |  | -104.8***  (-155.3, -54.4) |  | -177.6***  (-248.3, -106.9) |  | -6.2***  (-7.8, -4.6) |
| AIC | 133496786 | 133495757 | 80892311 | 80890431 | -63897 | -66395 |
| BIC | 134755488 | 134754487 | 81814531 | 81812678 | 1194805 | 1192334 |

**Supplementary Table 12:** Model estimate comparison table for pay overall, extending the baseline period (* = p value <0.05, ** = p value <0.01 and *** = p value < 0.001)

| **Time to/since surgery (months)** | **Main model (£)** | **Baseline period 12 months** (£) | **Baseline period 24 months** (£) |
| --- | --- | --- | --- |
| -60 to -54 | -7.9 (-29.5, 13.8) | -7.0 (-27.9, 13.9) | -5.6 (-25.1, 14.0) |
| -54 to -48 | -4.9 (-24.4, 14.5) | -4.1 (-22.9, 14.7) | -2.6 (-20.0, 14.7) |
| -48 to -42 | -5.6 (-23.0, 11.7) | -4.8 (-21.4, 11.8) | -3.3 (-18.4, 11.7) |
| -42 to -36 | -4.5 (-19.9, 10.9) | -3.7 (-18.3, 10.9) | -2.3 (-15.2, 10.7) |
| -36 to -30 | -0.4 (-14.0, 13.3) | 0.5 (-12.2, 13.2) | 1.9 (-9.0, 12.7) |
| -30 to -24 | 0.5 (-11.2, 12.3) | 1.4 (-9.4, 12.1) | 2.8 (-5.7, 11.3) |
| -24 to -18 | -3.1 (-13.2, 7.0) | -2.3 (-11.0, 6.5) |  |
| -18 to -12 | -4.3 (-12.4, 3.8) | -3.5 (-10.0, 3.1) |  |
| -12 to -6 | -1.7 (-7.6, 4.3) |  |  |
| 0 to 1 | -52.9*** (-59.4, -46.4) | -52.1*** (-59.0, -45.2) | -50.8*** (-58.8, -42.8) |
| 1 to 2 | -116.9*** (-124.0, -109.8) | -116.1*** (-123.4, -108.7) | -114.8*** (-123.1, -106.5) |
| 2 to 3 | -59.7*** (-67.2, -52.2) | -58.9*** (-66.7, -51.1) | -57.6*** (-66.4, -48.9) |
| 3 to 4 | -19.8*** (-27.6, -12.0) | -19.0*** (-27.1, -10.9) | -17.7*** (-26.7, -8.7) |
| 4 to 5 | -2.4 (-10.5, 5.6) | -1.6 (-9.9, 6.7) | -0.4 (-9.6, 8.9) |
| 5 to 6 | 3.5 (-5.0, 12.0) | 4.3 (-4.5, 13.1) | 5.6 (-4.1, 15.3) |
| 6 to 12 | 24.2*** (16.2, 32.2) | 25.0*** (16.6, 33.4) | 26.3*** (16.9, 35.7) |
| 12 to 18 | 47.1*** (37.2, 57.0) | 47.9*** (37.7, 58.0) | 49.1*** (38.1, 60.2) |
| 18 to 24 | 60.5*** (48.9, 72.1) | 61.3*** (49.4, 73.2) | 62.5*** (49.8, 75.3) |
| 24 to 30 | 63.9*** (50.8, 77.1) | 64.7*** (51.3, 78.2) | 66.0*** (51.6, 80.3) |
| 30 to 36 | 63.2*** (48.5, 77.8) | 64.0*** (49.0, 79.0) | 65.2*** (49.4, 81.0) |
| 36 to 42 | 70.8*** (54.6, 87.0) | 71.6*** (55.1, 88.1) | 72.8*** (55.4, 90.1) |
| 42 to 48 | 78.7*** (60.7, 96.7) | 79.5*** (61.1, 97.9) | 80.7*** (61.5, 99.8) |
| 48 to 54 | 78.6*** (58.9, 98.3) | 79.4*** (59.4, 99.5) | 80.6*** (59.7, 101.4) |
| 54 to 60 | 84.4*** (63.1, 105.8) | 85.2*** (63.5, 106.9) | 86.4*** (63.9, 108.8) |
| AIC | 133496786 | 133496784 | 133496782 |
| BIC | 134755488 | 134755472 | 134755442 |

**Supplementary Table 13:** Model estimate comparison table for pay among those in work, extending the baseline period (* = p value <0.05, ** = p value <0.01 and *** = p value < 0.001)

| **Time to/since surgery (months)** | **Main model (£)** | **Baseline period 12_months (£)** | **Baseline period 24_months (£)** |
| --- | --- | --- | --- |
| -60 to -54 | 14.4 (-10.8, 39.6) | 16.1 (-8.1, 40.3) | 17.7 (-4.8, 40.2) |
| -54 to -48 | 5.0 (-18.3, 28.3) | 6.6 (-15.7, 29.0) | 8.2 (-12.3, 28.7) |
| -48 to -42 | -2.0 (-23.0, 19.0) | -0.4 (-20.3, 19.5) | 1.2 (-16.8, 19.2) |
| -42 to -36 | -2.9 (-21.5, 15.8) | -1.2 (-18.8, 16.3) | 0.3 (-15.2, 15.8) |
| -36 to -30 | -4.6 (-21.5, 12.3) | -2.9 (-18.5, 12.6) | -1.4 (-14.7, 11.9) |
| -30 to -24 | -3.7 (-18.6, 11.2) | -2.1 (-15.8, 11.6) | -0.5 (-11.5, 10.4) |
| -24 to -18 | -6.0 (-19.3, 7.2) | -4.4 (-15.9, 7.1) |  |
| -18 to -12 | -3.3 (-14.1, 7.5) | -1.7 (-10.6, 7.2) |  |
| -12 to -6 | -3.3 (-12.1, 5.5) |  |  |
| 0 to 1 | -86.3*** (-97.4, -75.1) | -84.7*** (-95.9, -73.4) | -83.3*** (-95.4, -71.3) |
| 1 to 2 | -206.0*** (-217.8, -194.2) | -204.4*** (-216.1, -192.7) | -203.1*** (-215.5, -190.7) |
| 2 to 3 | -106.4*** (-118.7, -94.2) | -104.9*** (-117.0, -92.7) | -103.5*** (-116.4, -90.7) |
| 3 to 4 | -42.1*** (-54.3, -29.9) | -40.5*** (-52.7, -28.3) | -39.2*** (-52.1, -26.3) |
| 4 to 5 | -17.6** (-30.0, -5.1) | -16.0* (-28.3, -3.7) | -14.6* (-27.7, -1.6) |
| 5 to 6 | -10.1 (-23.2, 3.0) | -8.5 (-21.5, 4.5) | -7.2 (-20.9, 6.5) |
| 6 to 12 | 5.9 (-5.0, 16.8) | 7.5 (-3.5, 18.5) | 8.8 (-3.1, 20.7) |
| 12 to 18 | 18.8** (5.5, 32.2) | 20.4** (7.2, 33.6) | 21.7** (7.8, 35.6) |
| 18 to 24 | 21.0** (6.0, 35.9) | 22.5** (7.5, 37.6) | 23.8** (8.1, 39.6) |
| 24 to 30 | 14.9 (-2.0, 31.9) | 16.5 (-0.5, 33.6) | 17.8* (0.0, 35.5) |
| 30 to 36 | 8.3 (-10.0, 26.7) | 9.9 (-8.6, 28.5) | 11.1 (-8.1, 30.4) |
| 36 to 42 | 14.1 (-5.9, 34.1) | 15.7 (-4.5, 35.8) | 16.9 (-4.0, 37.7) |
| 42 to 48 | 16.3 (-5.8, 38.5) | 17.9 (-4.5, 40.3) | 19.1 (-4.0, 42.2) |
| 48 to 54 | 17.5 (-6.6, 41.6) | 19.0 (-5.3, 43.4) | 20.2 (-4.8, 45.2) |
| 54 to 60 | 17.4 (-8.8, 43.5) | 18.9 (-7.5, 45.3) | 20.1 (-7.0, 47.1) |
| AIC | 80892311 | 80892310 | 80892307 |
| BIC | 81814531 | 81814516 | 81814487 |

**Supplementary Table 14:** Model estimate comparison table for probability of employment, extending the baseline period (* = p value <0.05, ** = p value <0.01 and *** = p value < 0.001)

| **Time to/since surgery (months)** | **Main model (percentage points)** | **Baseline period 12 months (percentage points)** | **Baseline period 24 months (percentage points)** |
| --- | --- | --- | --- |
| -60 to -54 | 1.7*** (1.0, 2.3) | 1.6*** (1.0, 2.2) | 1.5*** (0.9, 2.1) |
| -54 to -48 | 1.5*** (0.9, 2.1) | 1.5*** (0.9, 2.0) | 1.3*** (0.8, 1.8) |
| -48 to -42 | 1.4*** (0.9, 1.9) | 1.4*** (0.9, 1.9) | 1.2*** (0.8, 1.7) |
| -42 to -36 | 1.3*** (0.8, 1.7) | 1.2*** (0.8, 1.7) | 1.1*** (0.7, 1.5) |
| -36 to -30 | 1.1*** (0.7, 1.5) | 1.1*** (0.7, 1.5) | 0.9*** (0.6, 1.3) |
| -30 to -24 | 0.7*** (0.3, 1.1) | 0.7*** (0.3, 1.0) | 0.5*** (0.3, 0.8) |
| -24 to -18 | 0.4* (0.1, 0.7) | 0.3* (0.1, 0.6) |  |
| -18 to -12 | 0.2 (-0.0, 0.5) | 0.2 (-0.0, 0.4) |  |
| -12 to -6 | 0.1 (-0.1, 0.3) |  |  |
| 0 to 1 | -0.6*** (-0.8, -0.5) | -0.7*** (-0.8, -0.5) | -0.8*** (-1.0, -0.6) |
| 1 to 2 | -0.8*** (-1.0, -0.6) | -0.9*** (-1.0, -0.7) | -1.0*** (-1.2, -0.7) |
| 2 to 3 | -0.4*** (-0.6, -0.2) | -0.4*** (-0.6, -0.2) | -0.5*** (-0.8, -0.3) |
| 3 to 4 | 0.1 (-0.1, 0.3) | 0.0 (-0.2, 0.2) | -0.1 (-0.4, 0.1) |
| 4 to 5 | 0.5*** (0.2, 0.7) | 0.4*** (0.2, 0.7) | 0.3* (0.0, 0.6) |
| 5 to 6 | 0.7*** (0.4, 0.9) | 0.6*** (0.4, 0.9) | 0.5*** (0.2, 0.8) |
| 6 to 12 | 1.5*** (1.3, 1.7) | 1.5*** (1.2, 1.7) | 1.3*** (1.1, 1.6) |
| 12 to 18 | 2.5*** (2.2, 2.8) | 2.4*** (2.1, 2.7) | 2.3*** (2.0, 2.6) |
| 18 to 24 | 3.1*** (2.8, 3.5) | 3.1*** (2.7, 3.4) | 3.0*** (2.6, 3.3) |
| 24 to 30 | 3.5*** (3.1, 3.9) | 3.4*** (3.0, 3.8) | 3.3*** (2.9, 3.7) |
| 30 to 36 | 3.7*** (3.2, 4.1) | 3.6*** (3.2, 4.0) | 3.5*** (3.1, 3.9) |
| 36 to 42 | 3.9*** (3.4, 4.4) | 3.9*** (3.4, 4.3) | 3.8*** (3.3, 4.2) |
| 42 to 48 | 4.2*** (3.7, 4.7) | 4.2*** (3.7, 4.7) | 4.1*** (3.6, 4.6) |
| 48 to 54 | 4.2*** (3.7, 4.7) | 4.1*** (3.6, 4.7) | 4.0*** (3.5, 4.6) |
| 54 to 60 | 4.3*** (3.7, 4.9) | 4.3*** (3.7, 4.9) | 4.2*** (3.6, 4.8) |
| AIC | -63897 | -63897 | -63871 |
| BIC | 1194805 | 1194791 | 1194789 |

**Supplementary Table 15:** Model estimate comparison table for pay overall, changing the baseline period timing to an earlier pre-surgery period (* = p value <0.05, ** = p value <0.01 and *** = p value < 0.001)

| **Time to/since surgery (months)** | **Main model (£)** | **Baseline period 6 months earlier (£)** | **Baseline period 12 months earlier (£)** | **Baseline period 24 months earlier (£)** |
| --- | --- | --- | --- | --- |
| -60 to -54 | -7.9  (-29.5, 13.8) | -6.2  (-26.8, 14.4) | -3.6  (-23.2, 16.1) | -8.4  (-25.7, 8.9) |
| -54 to -48 | -4.9  (-24.4, 14.5) | -3.2  (-21.8, 15.3) | -0.6  (-18.1, 16.8) | -5.5  (-20.3, 9.4) |
| -48 to -42 | -5.6  (-23.0, 11.7) | -3.9  (-20.3, 12.4) | -1.3  (-16.6, 14.0) | -6.2  (-18.8, 6.4) |
| -42 to -36 | -4.5  (-19.9, 10.9) | -2.8  (-17.2, 11.6) | -0.2  (-13.4, 13.0) | -5.1  (-15.2, 5.0) |
| -36 to -30 | -0.4  (-14.0, 13.3) | 1.3  (-11.1, 13.7) | 3.9  (-7.4, 15.2) | -0.9  (-8.1, 6.3) |
| -30 to -24 | 0.5  (-11.2, 12.3) | 2.2  (-8.3, 12.7) | 4.8  (-4.0, 13.7) |  |
| -24 to -18 | -3.1  (-13.2, 7.0) | -1.4  (-9.7, 6.9) | 1.2  (-5.2, 7.6) | -3.6  (-10.3, 3.1) |
| -18 to -12 | -4.3  (-12.4, 3.8) | -2.6  (-8.7, 3.5) |  | -4.8  (-13.7, 4.0) |
| -12 to -6 | -1.7  (-7.6, 4.3) |  | 2.6  (-3.5, 8.7) | -2.2  (-12.7, 8.3) |
| -6 to 0 |  | 1.7  (-4.3, 7.6) | 4.3  (-3.8, 12.4) | -0.5  (-12.3, 11.2) |
| 0 to 1 | -52.9***  (-59.4, -46.4) | -51.2***  (-59.7, -42.7) | -48.6***  (-58.9, -38.4) | -53.5***  (-66.8, -40.1) |
| 1 to 2 | -116.9***  (-124.0, -109.8) | -115.2***  (-124.0, -106.4) | -112.6***  (-123.2, -102.0) | -117.4***  (-130.8, -104.1) |
| 2 to 3 | -59.7***  (-67.2, -52.2) | -58.0***  (-67.2, -48.9) | -55.4***  (-66.3, -44.5) | -60.3***  (-74.1, -46.4) |
| 3 to 4 | -19.8***  (-27.6, -12.0) | -18.1***  (-27.6, -8.7) | -15.5**  (-26.6, -4.4) | -20.4**  (-34.5, -6.3) |
| 4 to 5 | -2.4  (-10.5, 5.6) | -0.8  (-10.4, 8.9) | 1.8  (-9.5, 13.2) | -3.0  (-17.3, 11.3) |
| 5 to 6 | 3.5  (-5.0, 12.0) | 5.2  (-4.9, 15.3) | 7.8  (-3.9, 19.5) | 2.9  (-11.6, 17.5) |
| 6 to 12 | 24.2***  (16.2, 32.2) | 25.9***  (16.0, 35.7) | 28.5***  (17.0, 39.9) | 23.6**  (9.1, 38.1) |
| 12 to 18 | 47.1***  (37.2, 57.0) | 48.8***  (37.4, 60.1) | 51.4***  (38.4, 64.3) | 46.5***  (30.6, 62.5) |
| 18 to 24 | 60.5***  (48.9, 72.1) | 62.2***  (49.1, 75.2) | 64.8***  (50.3, 79.3) | 60.0***  (42.5, 77.4) |
| 24 to 30 | 63.9***  (50.8, 77.1) | 65.6***  (51.1, 80.1) | 68.2***  (52.3, 84.2) | 63.4***  (44.5, 82.2) |
| 30 to 36 | 63.2***  (48.5, 77.8) | 64.9***  (48.8, 80.9) | 67.5***  (50.1, 84.8) | 62.6***  (42.4, 82.9) |
| 36 to 42 | 70.8***  (54.6, 87.0) | 72.5***  (55.0, 89.9) | 75.1***  (56.2, 93.9) | 70.2***  (48.5, 91.9) |
| 42 to 48 | 78.7***  (60.7, 96.7) | 80.4***  (61.1, 99.7) | 83.0***  (62.4, 103.6) | 78.1***  (54.7, 101.6) |
| 48 to 54 | 78.6***  (58.9, 98.3) | 80.3***  (59.4, 101.3) | 82.9***  (60.6, 105.2) | 78.1***  (53.1, 103.1) |
| 54 to 60 | 84.4***  (63.1, 105.8) | 86.1***  (63.6, 108.7) | 88.7***  (64.9, 112.6) | 83.9***  (57.4, 110.4) |
| AIC | 133496786 | 133496786 | 133496786 | 133496786 |
| BIC | 134755488 | 134755488 | 134755488 | 134755488 |

**Supplementary Table 16:** Model estimate comparison table for pay among those in work, changing the baseline period timing to an earlier pre-surgery period (* = p value <0.05, ** = p value <0.01 and *** = p value < 0.001)

| **Time to/since surgery (months)** | **Main model (£)** | **Baseline period 6 months earlier (£)** | **Baseline period 12 months earlier (£)** | **Baseline period 24 months earlier (£)** |
| --- | --- | --- | --- | --- |
| -60 to -54 | 14.4  (-10.8, 39.6) | 17.7  (-6.2, 41.7) | 17.8  (-5.1, 40.7) | 18.1  (-2.6, 38.8) |
| -54 to -48 | 5.0  (-18.3, 28.3) | 8.3  (-14.0, 30.5) | 8.3  (-12.5, 29.2) | 8.7  (-9.4, 26.8) |
| -48 to -42 | -2.0  (-23.0, 19.0) | 1.3  (-18.4, 21.0) | 1.3  (-17.2, 19.9) | 1.7  (-14.1, 17.5) |
| -42 to -36 | -2.9  (-21.5, 15.8) | 0.4  (-17.1, 17.9) | 0.5  (-15.6, 16.5) | 0.8  (-12.0, 13.6) |
| -36 to -30 | -4.6  (-21.5, 12.3) | -1.3  (-16.7, 14.2) | -1.2  (-15.5, 13.0) | -0.9  (-10.8, 9.0) |
| -30 to -24 | -3.7  (-18.6, 11.2) | -0.4  (-14.2, 13.4) | -0.4  (-12.1, 11.4) |  |
| -24 to -18 | -6.0  (-19.3, 7.2) | -2.7  (-14.0, 8.5) | -2.7  (-12.0, 6.6) | -2.3  (-11.8, 7.1) |
| -18 to -12 | -3.3  (-14.1, 7.5) | -0.0  (-9.0, 8.9) |  | 0.4  (-11.4, 12.1) |
| -12 to -6 | -3.3  (-12.1, 5.5) |  | 0.0  (-8.9, 9.0) | 0.4  (-13.4, 14.2) |
| -6 to 0 |  | 3.3  (-5.5, 12.1) | 3.3  (-7.5, 14.1) | 3.7  (-11.2, 18.6) |
| 0 to 1 | -86.3***  (-97.4, -75.1) | -83.0***  (-96.0, -70.0) | -82.9***  (-97.5, -68.3) | -82.6***  (-100.4, -64.7) |
| 1 to 2 | -206.0***  (-217.8, -194.2) | -202.7***  (-215.9, -189.5) | -202.7***  (-217.7, -187.7) | -202.3***  (-220.1, -184.5) |
| 2 to 3 | -106.4***  (-118.7, -94.2) | -103.1***  (-116.8, -89.5) | -103.1***  (-118.4, -87.8) | -102.7***  (-121.1, -84.4) |
| 3 to 4 | -42.1***  (-54.3, -29.9) | -38.8***  (-52.5, -25.1) | -38.8***  (-54.2, -23.3) | -38.4***  (-57.0, -19.8) |
| 4 to 5 | -17.6**  (-30.0, -5.1) | -14.3*  (-28.0, -0.5) | -14.2  (-29.7, 1.3) | -13.9  (-32.5, 4.8) |
| 5 to 6 | -10.1  (-23.2, 3.0) | -6.8  (-21.2, 7.6) | -6.7  (-22.7, 9.2) | -6.4  (-25.5, 12.7) |
| 6 to 12 | 5.9  (-5.0, 16.8) | 9.2  (-3.6, 22.0) | 9.2  (-5.1, 23.6) | 9.6  (-8.3, 27.5) |
| 12 to 18 | 18.8**  (5.5, 32.2) | 22.1**  (7.5, 36.7) | 22.2**  (6.0, 38.4) | 22.5*  (2.9, 42.1) |
| 18 to 24 | 21.0**  (6.0, 35.9) | 24.3**  (7.7, 40.8) | 24.3**  (6.6, 42.0) | 24.7*  (3.5, 45.8) |
| 24 to 30 | 14.9  (-2.0, 31.9) | 18.2  (-0.1, 36.5) | 18.3  (-1.4, 38.0) | 18.6  (-4.3, 41.6) |
| 30 to 36 | 8.3  (-10.0, 26.7) | 11.6  (-8.2, 31.5) | 11.7  (-9.4, 32.7) | 12.0  (-12.3, 36.4) |
| 36 to 42 | 14.1  (-5.9, 34.1) | 17.4  (-3.9, 38.7) | 17.4  (-5.2, 40.1) | 17.8  (-8.1, 43.7) |
| 42 to 48 | 16.3  (-5.8, 38.5) | 19.6  (-3.9, 43.2) | 19.7  (-5.1, 44.4) | 20.0  (-7.9, 48.0) |
| 48 to 54 | 17.5  (-6.6, 41.6) | 20.8  (-4.6, 46.1) | 20.8  (-5.8, 47.5) | 21.2  (-8.5, 50.9) |
| 54 to 60 | 17.4  (-8.8, 43.5) | 20.7  (-6.8, 48.1) | 20.7  (-7.9, 49.4) | 21.1  (-10.5, 52.6) |
| AIC | 80892311 | 80892311 | 80892311 | 80892311 |
| BIC | 81814531 | 81814531 | 81814531 | 81814531 |

**Supplementary Table 17:** Model estimate comparison table for probability of employment, changing the baseline period timing to an earlier pre-surgery period (* = p value <0.05, ** = p value <0.01 and *** = p value < 0.001)

| **Time to/since surgery (months)** | **Main model (percentage points)** | **Baseline period 6 months earlier (percentage points)** | **Baseline period 12 months earlier (percentage points)** | **Baseline period 24 months earlier (percentage points)** |
| --- | --- | --- | --- | --- |
| -60 to -54 | 1.7*** (1.0, 2.3) | 1.6*** (1.0, 2.2) | 1.5*** (0.9, 2.1) | 1.0*** (0.4, 1.5) |
| -54 to -48 | 1.5*** (0.9, 2.1) | 1.4*** (0.9, 2.0) | 1.3*** (0.8, 1.8) | 0.8*** (0.3, 1.3) |
| -48 to -42 | 1.4*** (0.9, 1.9) | 1.3*** (0.8, 1.8) | 1.2*** (0.7, 1.7) | 0.7*** (0.3, 1.1) |
| -42 to -36 | 1.3*** (0.8, 1.7) | 1.2*** (0.7, 1.6) | 1.0*** (0.6, 1.5) | 0.6*** (0.2, 0.9) |
| -36 to -30 | 1.1*** (0.7, 1.5) | 1.0*** (0.6, 1.4) | 0.9*** (0.6, 1.3) | 0.4*** (0.2, 0.6) |
| -30 to -24 | 0.7*** (0.3, 1.1) | 0.6*** (0.3, 0.9) | 0.5*** (0.2, 0.8) |  |
| -24 to -18 | 0.4* (0.1, 0.7) | 0.3* (0.0, 0.6) | 0.2 (-0.0, 0.4) | -0.3** (-0.5, -0.1) |
| -18 to -12 | 0.2 (-0.0, 0.5) | 0.1 (-0.1, 0.3) |  | -0.5*** (-0.8, -0.2) |
| -12 to -6 | 0.1 (-0.1, 0.3) |  | -0.1 (-0.3, 0.1) | -0.6*** (-0.9, -0.3) |
| -6 to 0 |  | -0.1 (-0.3, 0.1) | -0.2 (-0.5, 0.0) | -0.7*** (-1.1, -0.3) |
| 0 to 1 | -0.6*** (-0.8, -0.5) | -0.7*** (-1.0, -0.5) | -0.8*** (-1.1, -0.6) | -1.3*** (-1.7, -0.9) |
| 1 to 2 | -0.8*** (-1.0, -0.6) | -0.9*** (-1.2, -0.7) | -1.0*** (-1.3, -0.7) | -1.5*** (-1.9, -1.1) |
| 2 to 3 | -0.4*** (-0.6, -0.2) | -0.5*** (-0.7, -0.2) | -0.6*** (-0.9, -0.3) | -1.1*** (-1.5, -0.7) |
| 3 to 4 | 0.1 (-0.1, 0.3) | -0.0 (-0.3, 0.2) | -0.2 (-0.5, 0.2) | -0.6** (-1.1, -0.2) |
| 4 to 5 | 0.5*** (0.2, 0.7) | 0.4** (0.1, 0.6) | 0.2 (-0.1, 0.6) | -0.2 (-0.6, 0.2) |
| 5 to 6 | 0.7*** (0.4, 0.9) | 0.6*** (0.3, 0.9) | 0.4** (0.1, 0.8) | -0.0 (-0.5, 0.4) |
| 6 to 12 | 1.5*** (1.3, 1.7) | 1.4*** (1.1, 1.7) | 1.3*** (0.9, 1.6) | 0.8*** (0.4, 1.2) |
| 12 to 18 | 2.5*** (2.2, 2.8) | 2.4*** (2.0, 2.7) | 2.2*** (1.9, 2.6) | 1.8*** (1.3, 2.2) |
| 18 to 24 | 3.1*** (2.8, 3.5) | 3.0*** (2.6, 3.4) | 2.9*** (2.5, 3.3) | 2.4*** (1.9, 2.9) |
| 24 to 30 | 3.5*** (3.1, 3.9) | 3.4*** (3.0, 3.8) | 3.2*** (2.8, 3.7) | 2.8*** (2.2, 3.3) |
| 30 to 36 | 3.7*** (3.2, 4.1) | 3.6*** (3.1, 4.0) | 3.4*** (3.0, 3.9) | 3.0*** (2.4, 3.5) |
| 36 to 42 | 3.9*** (3.4, 4.4) | 3.8*** (3.3, 4.3) | 3.7*** (3.2, 4.2) | 3.2*** (2.6, 3.8) |
| 42 to 48 | 4.2*** (3.7, 4.7) | 4.1*** (3.6, 4.7) | 4.0*** (3.5, 4.6) | 3.5*** (2.9, 4.2) |
| 48 to 54 | 4.2*** (3.7, 4.7) | 4.1*** (3.5, 4.7) | 4.0*** (3.4, 4.6) | 3.5*** (2.8, 4.1) |
| 54 to 60 | 4.3*** (3.7, 4.9) | 4.2*** (3.6, 4.8) | 4.1*** (3.5, 4.7) | 3.6*** (2.9, 4.3) |
| AIC | -63897 | -63897 | -63897 | -63897 |
| BIC | 1194805 | 1194805 | 1194805 | 1194805 |

**Supplementary Table 18:** Model estimate comparison table for omitting COVID-19 pandemic period (* = p value <0.05, ** = p value <0.01 and *** = p value < 0.001)

|  | **Pay overall** | | **Pay among those in work** | | **Probability of employment** | |
| --- | --- | --- | --- | --- | --- | --- |
| **Time to/since surgery (months)** | **Main model** | **Omitting COVID-19** | **Main model** | **Omitting COVID-19** | **Main model** | **Omitting COVID-19** |
| -60 to -54 | -7.9  (-29.5, 13.8) | -1.8  (-25.4, 21.8) | 14.4  (-10.8, 39.6) | 37.1**  (9.9, 64.3) | 1.7***  (1.0, 2.3) | 1.3***  (0.6, 2.0) |
| -54 to -48 | -4.9  (-24.4, 14.5) | -0.0  (-21.6, 21.5) | 5.0  (-18.3, 28.3) | 28.7*  (3.2, 54.2) | 1.5***  (0.9, 2.1) | 1.1***  (0.5, 1.8) |
| -48 to -42 | -5.6  (-23.0, 11.7) | -0.1  (-19.4, 19.3) | -2.0  (-23.0, 19.0) | 18.9  (-4.3, 42.1) | 1.4***  (0.9, 1.9) | 1.1***  (0.5, 1.7) |
| -42 to -36 | -4.5  (-19.9, 10.9) | -0.1  (-17.5, 17.2) | -2.9  (-21.5, 15.8) | 13.5  (-7.3, 34.3) | 1.3***  (0.8, 1.7) | 1.0***  (0.5, 1.5) |
| -36 to -30 | -0.4  (-14.0, 13.3) | 3.6  (-12.0, 19.2) | -4.6  (-21.5, 12.3) | 10.4  (-8.7, 29.5) | 1.1***  (0.7, 1.5) | 0.9***  (0.4, 1.4) |
| -30 to -24 | 0.5  (-11.2, 12.3) | 4.0  (-9.6, 17.6) | -3.7  (-18.6, 11.2) | 10.2  (-6.8, 27.2) | 0.7***  (0.3, 1.1) | 0.5*  (0.1, 0.9) |
| -24 to -18 | -3.1  (-13.2, 7.0) | -0.9  (-12.5, 10.8) | -6.0  (-19.3, 7.2) | 4.7  (-10.4, 19.8) | 0.4*  (0.1, 0.7) | 0.2  (-0.1, 0.6) |
| -18 to -12 | -4.3  (-12.4, 3.8) | -2.7  (-12.0, 6.7) | -3.3  (-14.1, 7.5) | 5.6  (-6.7, 17.9) | 0.2  (-0.0, 0.5) | 0.1  (-0.2, 0.4) |
| -12 to -6 | -1.7  (-7.6, 4.3) | 0.4  (-6.6, 7.3) | -3.3  (-12.1, 5.5) | 2.2  (-7.9, 12.3) | 0.1  (-0.1, 0.3) | 0.1  (-0.1, 0.3) |
| 0 to 1 | -52.9***  (-59.4, -46.4) | -50.9***  (-58.3, -43.6) | -86.3***  (-97.4, -75.1) | -86.5***  (-99.2, -73.7) | -0.6***  (-0.8, -0.5) | -0.6***  (-0.8, -0.4) |
| 1 to 2 | -116.9***  (-124.0, -109.8) | -117.3***  (-125.2, -109.3) | -206.0***  (-217.8, -194.2) | -212.1***  (-225.5, -198.8) | -0.8***  (-1.0, -0.6) | -0.8***  (-1.0, -0.6) |
| 2 to 3 | -59.7***  (-67.2, -52.2) | -62.4***  (-70.7, -54.0) | -106.4***  (-118.7, -94.2) | -116.5***  (-130.1, -102.9) | -0.4***  (-0.6, -0.2) | -0.3**  (-0.6, -0.1) |
| 3 to 4 | -19.8***  (-27.6, -12.0) | -21.1***  (-29.9, -12.4) | -42.1***  (-54.3, -29.9) | -47.7***  (-61.4, -34.0) | 0.1  (-0.1, 0.3) | 0.1  (-0.2, 0.3) |
| 4 to 5 | -2.4  (-10.5, 5.6) | -5.0  (-13.9, 3.9) | -17.6**  (-30.0, -5.1) | -28.0***  (-41.6, -14.3) | 0.5***  (0.2, 0.7) | 0.5***  (0.3, 0.8) |
| 5 to 6 | 3.5  (-5.0, 12.0) | 4.1  (-5.5, 13.8) | -10.1  (-23.2, 3.0) | -17.2*  (-32.1, -2.3) | 0.7***  (0.4, 0.9) | 0.8***  (0.5, 1.1) |
| 6 to 12 | 24.2***  (16.2, 32.2) | 21.9***  (12.7, 31.0) | 5.9  (-5.0, 16.8) | -4.8  (-17.1, 7.5) | 1.5***  (1.3, 1.7) | 1.7***  (1.4, 1.9) |
| 12 to 18 | 47.1***  (37.2, 57.0) | 49.6***  (37.9, 61.2) | 18.8**  (5.5, 32.2) | 9.8  (-5.8, 25.4) | 2.5***  (2.2, 2.8) | 2.9***  (2.5, 3.3) |
| 18 to 24 | 60.5***  (48.9, 72.1) | 66.7***  (52.8, 80.6) | 21.0**  (6.0, 35.9) | 13.3  (-4.4, 31.0) | 3.1***  (2.8, 3.5) | 3.7***  (3.3, 4.1) |
| 24 to 30 | 63.9***  (50.8, 77.1) | 69.0***  (52.7, 85.2) | 14.9  (-2.0, 31.9) | 1.9  (-19.0, 22.7) | 3.5***  (3.1, 3.9) | 4.1***  (3.6, 4.6) |
| 30 to 36 | 63.2***  (48.5, 77.8) | 74.3***  (55.9, 92.7) | 8.3  (-10.0, 26.7) | -1.2  (-24.1, 21.8) | 3.7***  (3.2, 4.1) | 4.4***  (3.9, 5.0) |
| 36 to 42 | 70.8***  (54.6, 87.0) | 88.7***  (68.1, 109.4) | 14.1  (-5.9, 34.1) | 2.3  (-23.5, 28.1) | 3.9***  (3.4, 4.4) | 5.0***  (4.4, 5.6) |
| 42 to 48 | 78.7***  (60.7, 96.7) | 96.8***  (73.2, 120.3) | 16.3  (-5.8, 38.5) | 5.1  (-24.4, 34.5) | 4.2***  (3.7, 4.7) | 5.3***  (4.6, 6.0) |
| 48 to 54 | 78.6***  (58.9, 98.3) | 98.4***  (71.7, 125.1) | 17.5  (-6.6, 41.6) | 11.3  (-21.6, 44.2) | 4.2***  (3.7, 4.7) | 5.2***  (4.4, 6.0) |
| 54 to 60 | 84.4***  (63.1, 105.8) | 102.3***  (72.3, 132.2) | 17.4  (-8.8, 43.5) | -0.5  (-36.9, 35.9) | 4.3***  (3.7, 4.9) | 5.6***  (4.7, 6.5) |
| AIC | 133496786 | 96166071 | 80892311 | 58590530 | -63897 | -745497 |
| BIC | 134755488 | 97395155 | 81814531 | 59470488 | 1194805 | 483587 |

### Supplementary Text

**Supplementary Text 1:** Assignment of ‘operation date’ to individuals in the unexposed datasets

We randomly assigned an index date to individuals who did not undergo bariatric surgery, in the same proportions by calendar month as the number of operations each month for the individuals who underwent bariatric surgery and had an obesity diagnosis. The age on the index date can then be calculated. We then used stratified sampling to sample the dataset of individuals who did not undergo bariatric surgery in sex and five-year age group in the same proportions as for the individuals who underwent bariatric surgery and had an obesity diagnosis. **Supplementary Text 2:** Equation for modelling

$$Y_{it}=\alpha_{i}+\lambda_{t}+\beta_{1}T+\beta_{2}f({Age}_{it})$$

where $Y_{it}$ is the outcome (monthly employee pay or paid employment status), which varies for individuals ($i$) and over time (t)), $\alpha_{i}$ are individual level fixed effects, $\lambda_{t}$ are calendar time fixed effects, $T$ is time to/since surgery and $Age$ is age in years. Age was modelled as a natural cubic spline with four internal knots at the 20^th^, 40^th^, 60^th^ and 80^th^ centiles of its distribution and boundary knots at the 10^th^ and 90^th^ centiles of the age distribution, and coefficients $\beta_{2}$. The effect of time to/since surgery on the outcomes is given by the coefficients $\beta_{1}$.

**Supplementary Text 3:** Sensitivity tests

We investigated the extent to which the baseline period specification (six months prior to the month of surgery in the main analysis) affected the results by varying the timing and length of the baseline period.

Extending the baseline period from 6 months to 12 or 24 months before surgery increased slightly the estimates for employee pay, particularly for pay among those in work, but decreased the estimates of probabilities of being a paid employee (**Supplementary Tables 12-14**). Changes in estimates were within the confidence limits of the main analysis estimates. Moving the six-month baseline period to earlier time periods before surgery (6-12 months, 12-18 months, and 24-30 months before surgery) sequentially reduced the estimated post-surgery rise in the probability of being a paid employee, with estimates outside the confidence limits of the main results (**Supplementary Tables 15-17**). Among those in work, there was little change to employee pay after the initial decrease in pay following surgery, and there was a slight increase in the estimates for employee pay overall when moving the baseline period to 6-12 months and 12-24 months before surgery. However, there was a slight decrease when moving it to 24-30 months before surgery, all within the confidence limits of the main analysis estimates.

The COVID-19 pandemic caused changes to macro-level average pay and employment rates, as well as to the scheduling of surgeries (with a reduced number of elective procedures taking place during the public health emergency). We therefore investigated the extent to which controlling for calendar time addressed these changes in the main analysis by re-running the model while omitting months from March 2020 onwards.

Omitting data from the COVID-19 pandemic period resulted in consistently higher estimates of the association of surgery with the probability of being a paid employee, starting from 12 months after surgery (**Supplementary Table 18**). Higher estimates were also seen for employee pay overall, although within the confidence limits of the main analysis.

**Supplementary Text 4:** Placebo tests

We first tested for effects before surgery by censoring the data for the exposed cohort at the date of surgery using the period 30-36 months before surgery as the reference period, as well as all time-periods for the unexposed sample~~,~~ as in the main analysis.

We also tested for effects in the unexposed population by using only this sample, randomly assigning individuals in this sample to be either ‘treated’ or ‘untreated’, and then using the randomly assigned index date to set a reference period and time periods before/after surgery for the ‘treated’ group.

No effects were found for employee pay overall or pay among those in work for either placebo test (**Supplementary Figures 20** and **21**). For the first placebo test, censoring the data at index date and setting the reference period to be 36 to 30 months before this date, there was a small negative change in probability of being a paid employee after surgery, which is in line with the pre-surgery trends seen for this outcome. For the second placebo test, using only the unexposed data and randomly ‘treating’ half of this population, no effects were found for the probability of being a paid employee.

**Supplementary Text 4:** Unadjusted trends on outcomes in calendar time and by age and sex

Pay trends across calendar time were also broadly parallel for people who underwent bariatric surgery and those who did not apart from a slight drop from approximately April 2022 onwards for people who had undergone bariatric surgery, before their surgery took place (**Supplementary Figure 3**). However, employment trends increased overall over time for the people who underwent bariatric surgery, more so when omitting post-operation data, whereas they decreased overall for the people who did not undergo bariatric surgery. Both groups had a small drop in the probability of employment around April 2020, the first COVID-19 pandemic wave.

Trends in mean monthly deflated employee pay and the probability of being a paid employee differed slightly between the people who underwent bariatric surgery and those who did not by age and sex (**Supplementary Figure 4**). There was generally a larger difference in pay in midlife (ages ~40-50), with higher employee pay among the people who did not undergo bariatric surgery. The probability of being a paid employee increased from age 21 to 25 years among the people who did not undergo bariatric surgery but remained at a stable level for the exposed sample and started to decrease earlier among the people who underwent bariatric surgery (~ age 50 compared to ~age 54).

#### Supplementary Figures

**Supplementary Figure 1:** Dataset linkage and sample selection process

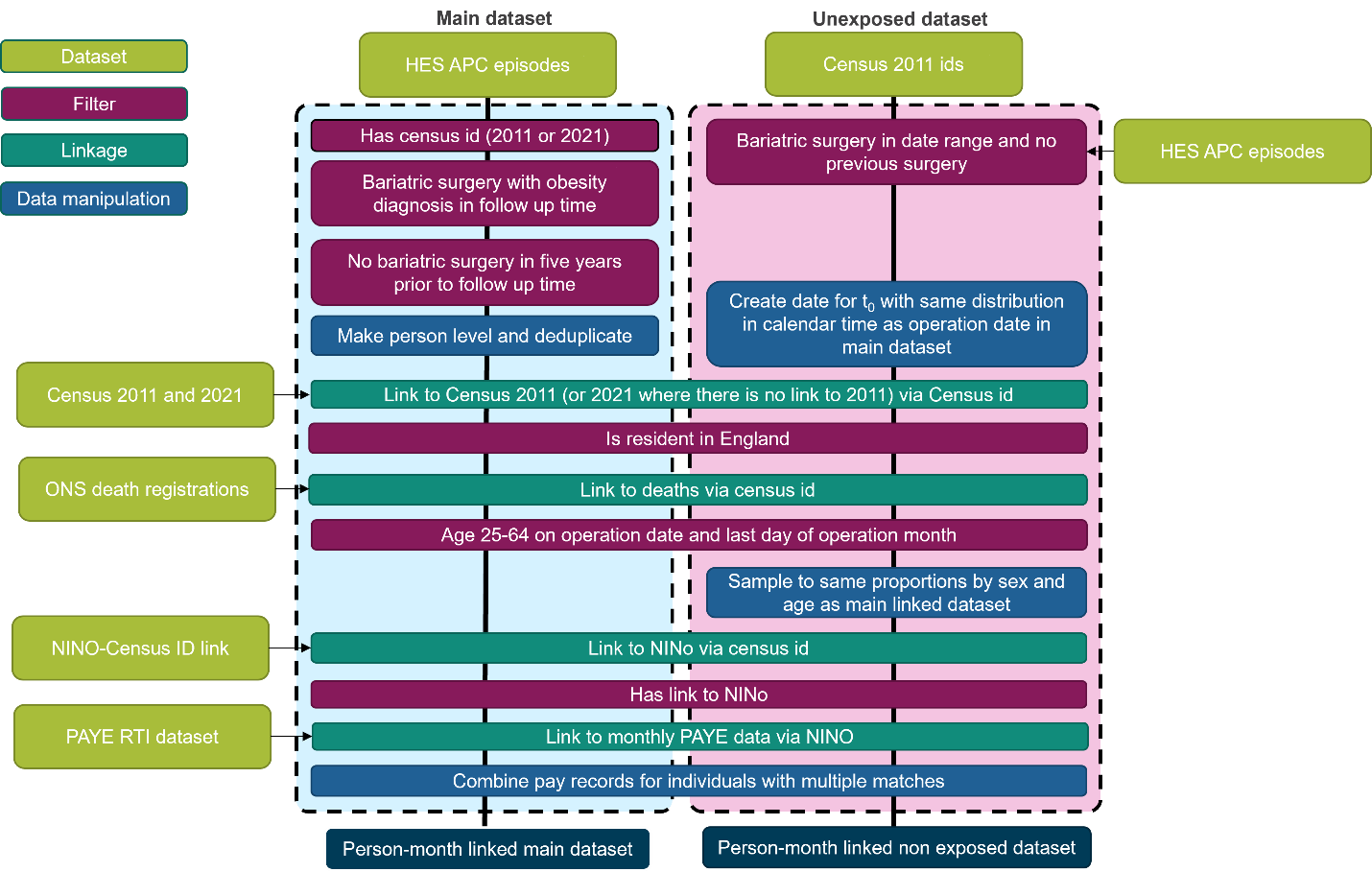

**Supplementary Figure 2:** Number of bariatric surgery operations, total and by type, over calendar time

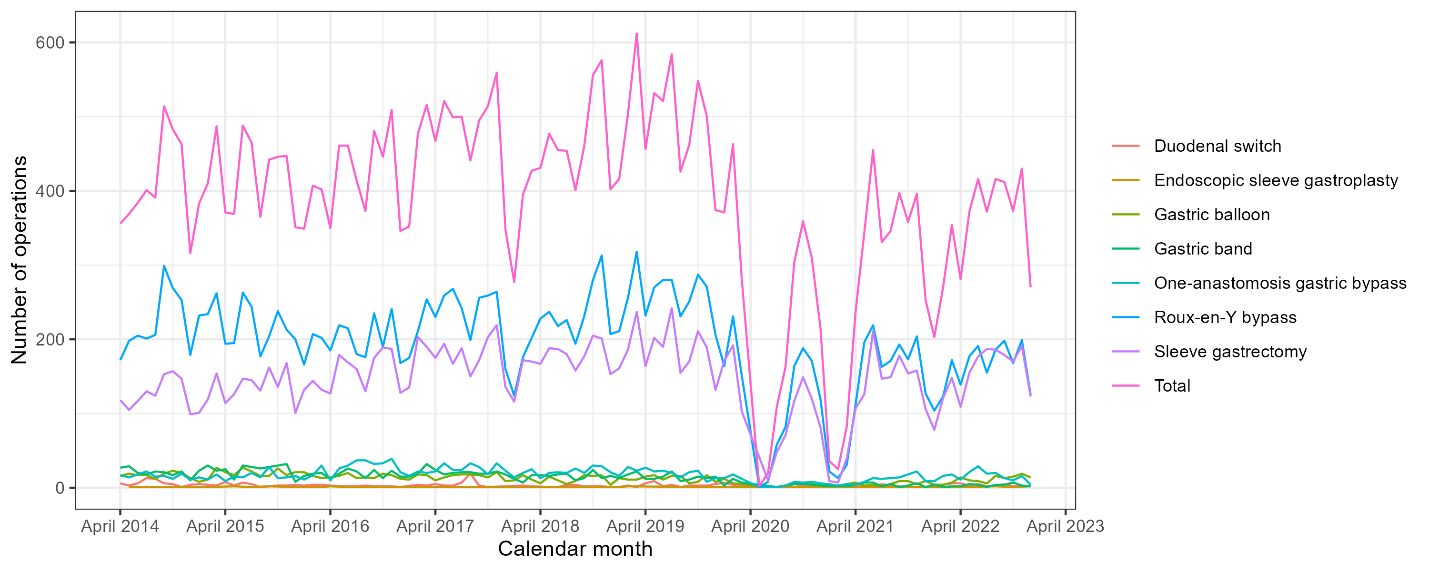

**Supplementary Figure 3:** Mean monthly pay and employment in calendar time

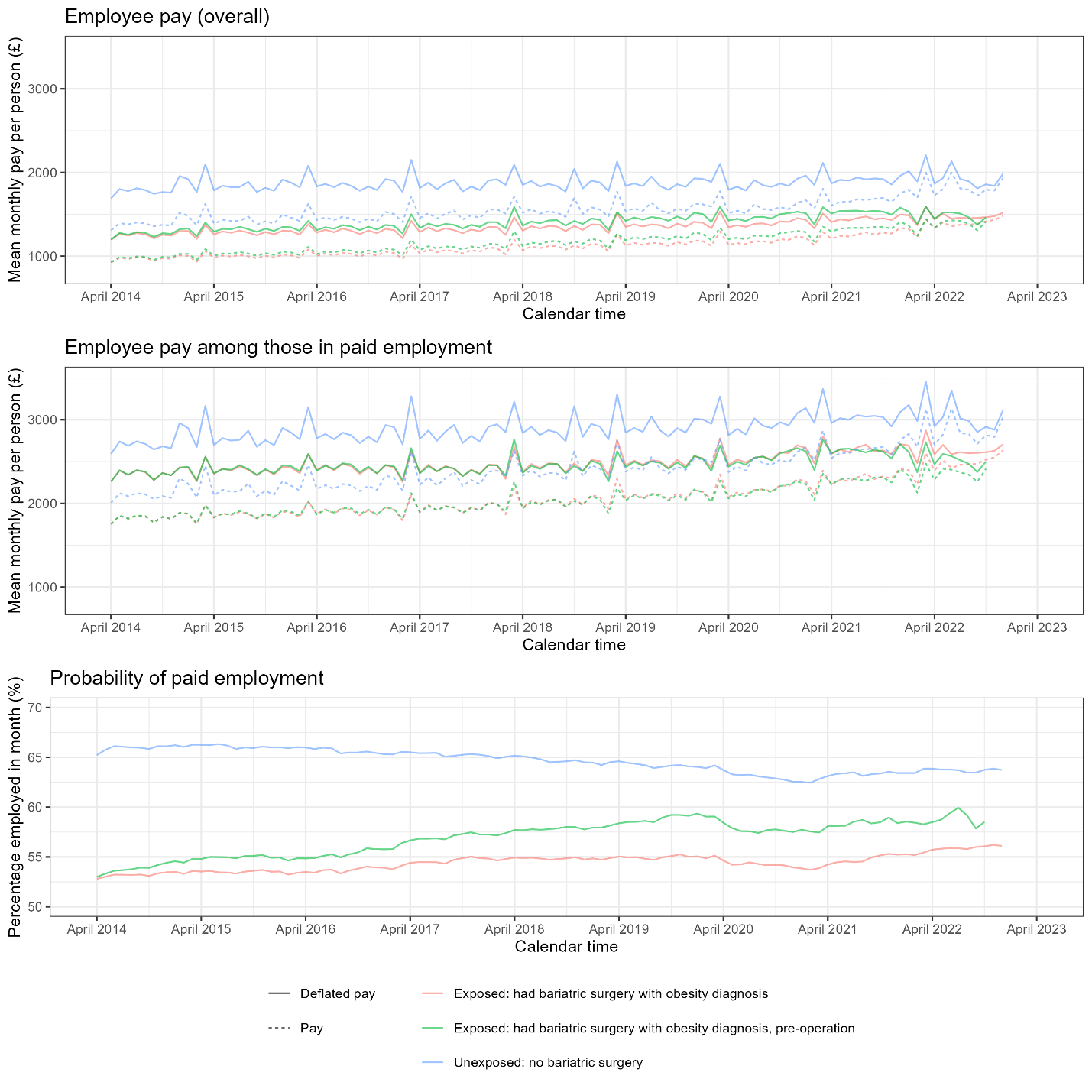

**Supplementary Figure 4:** Mean monthly pay and employment by age and sex

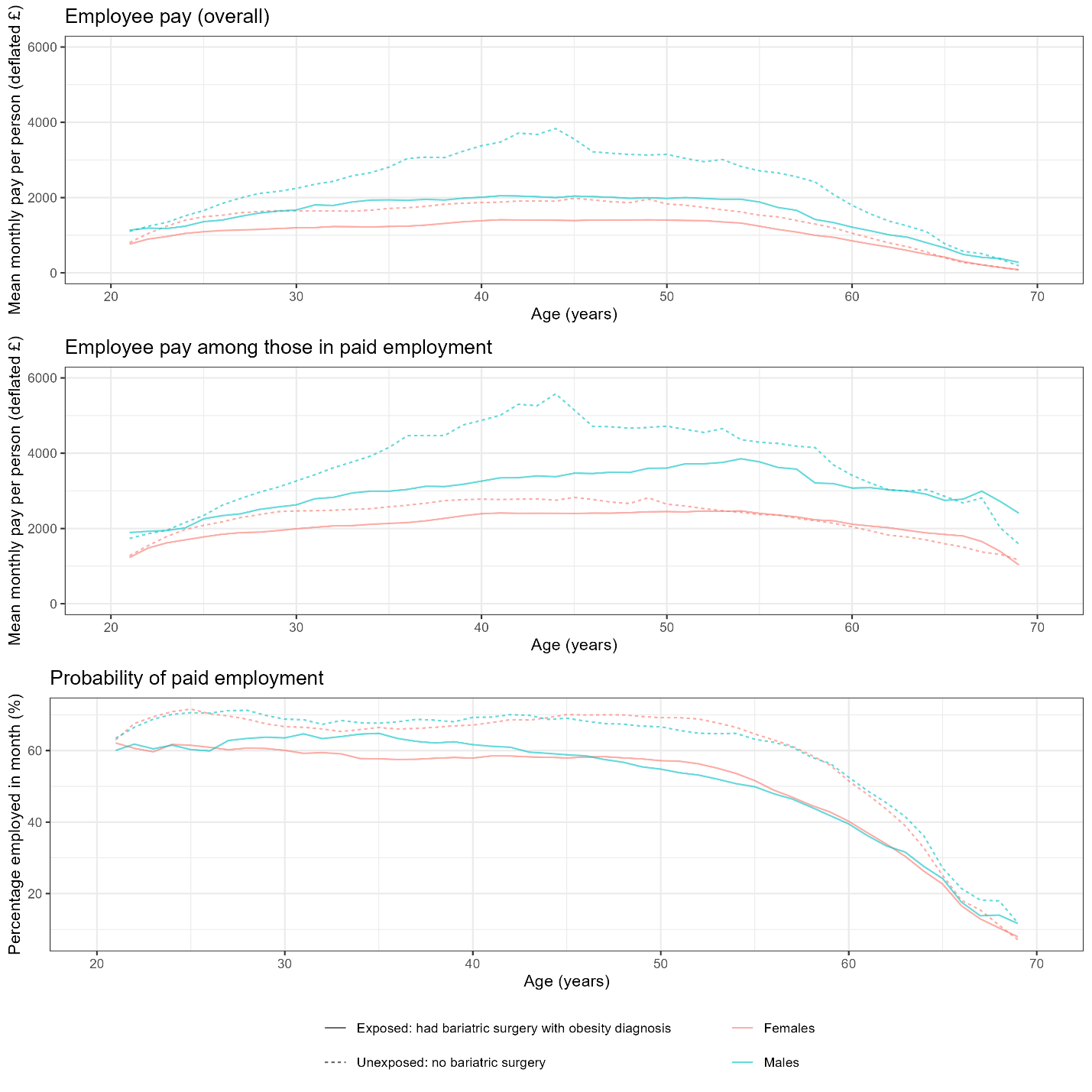

**Supplementary Figure 5:** Effect of bariatric surgery on monthly employee pay (overall) before and after surgery, across age groups

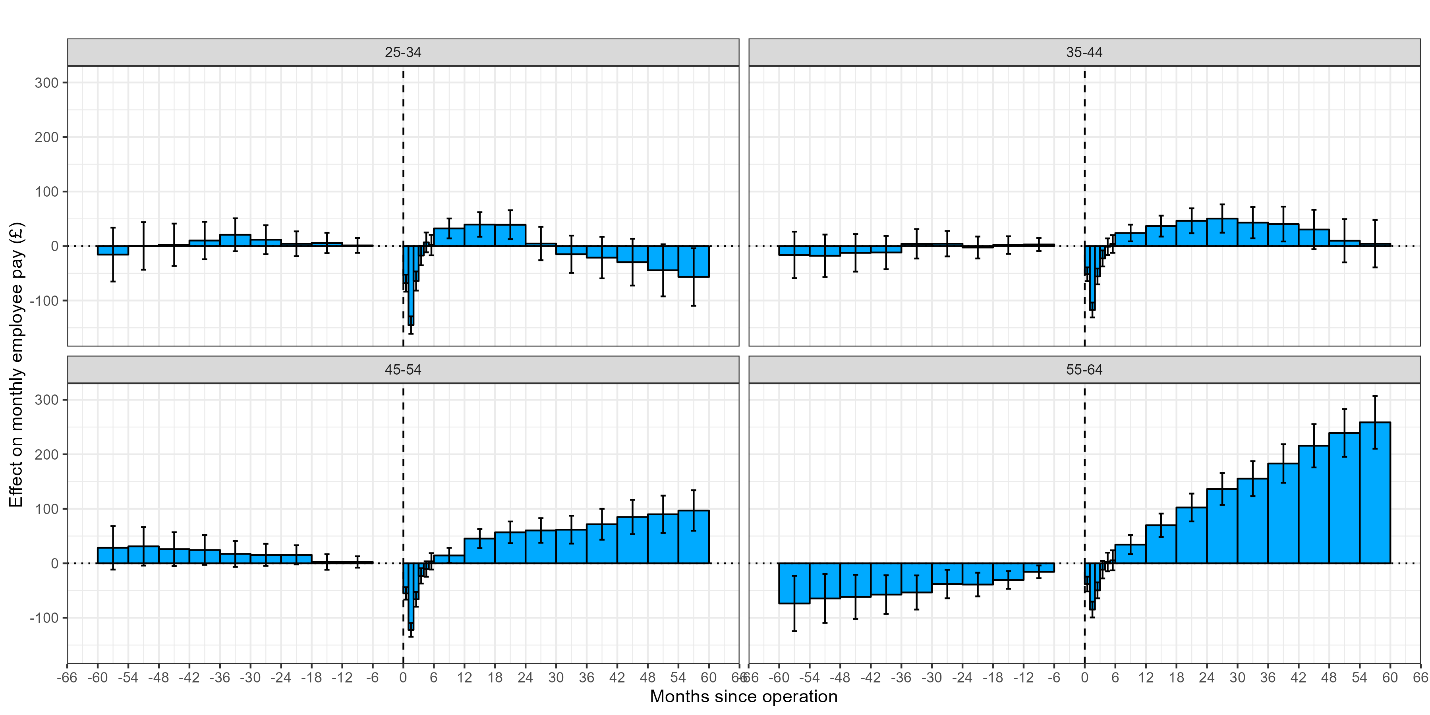

**Supplementary Figure 6:** Effect of bariatric surgery on monthly employee pay among those in work before and after surgery, across age groups

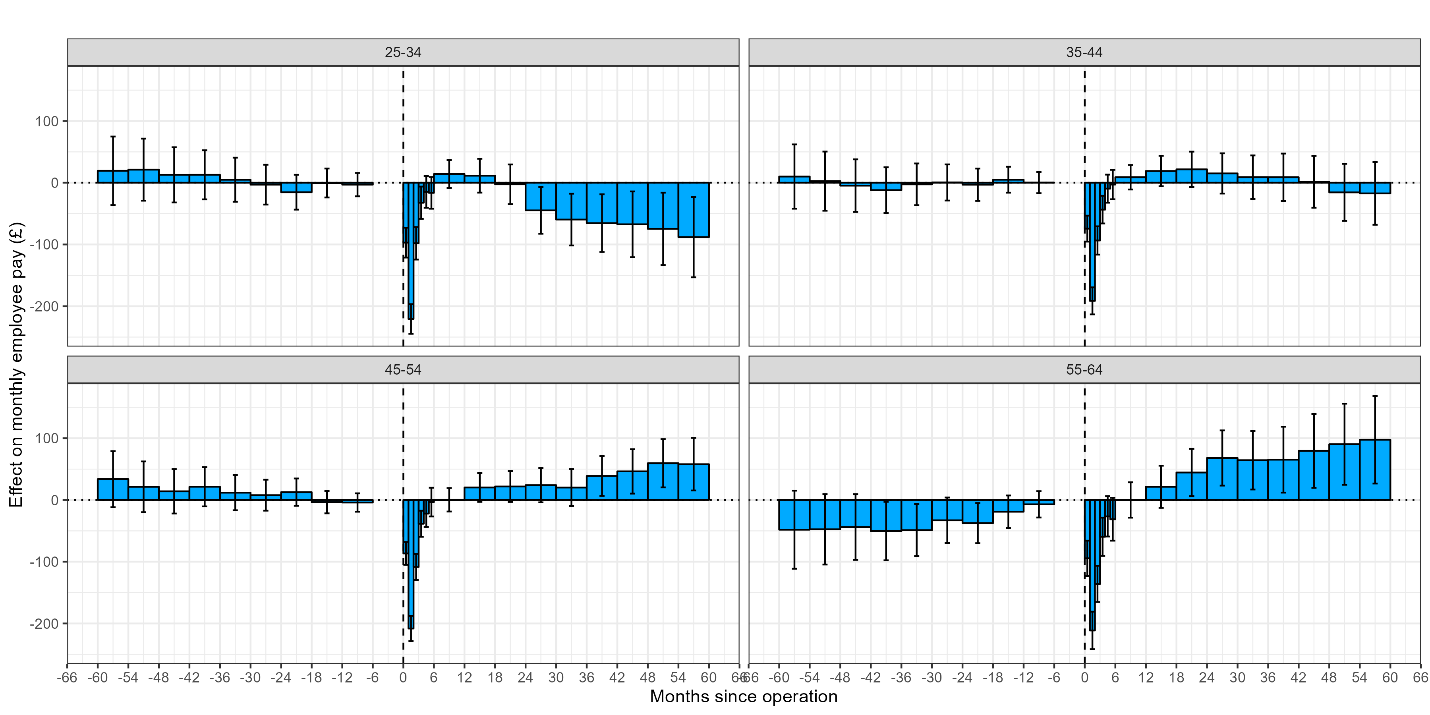

**Supplementary Figure 7:** Effect of bariatric surgery on probability of being a paid employee before and after surgery, across age groups

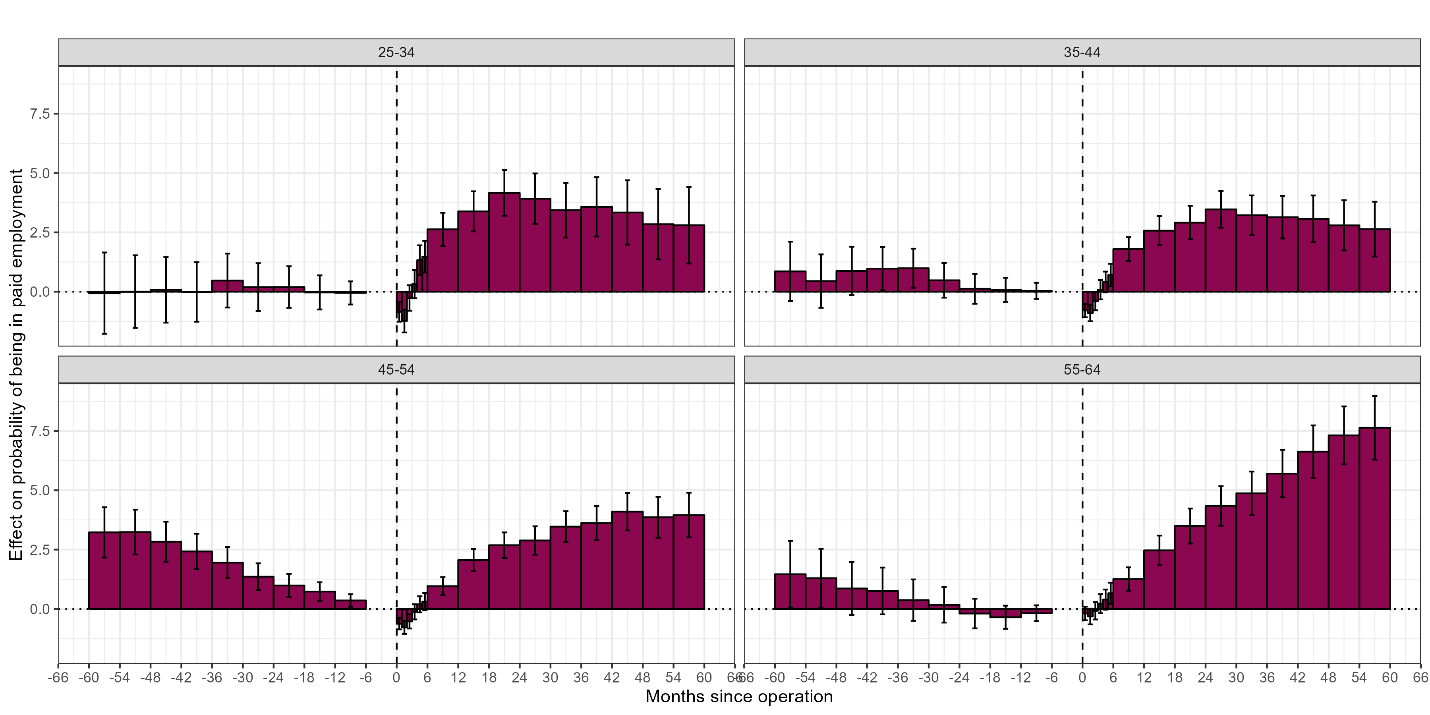

**Supplementary Figure 8:** Effect of bariatric surgery on monthly employee pay (overall) before and after surgery, by sex

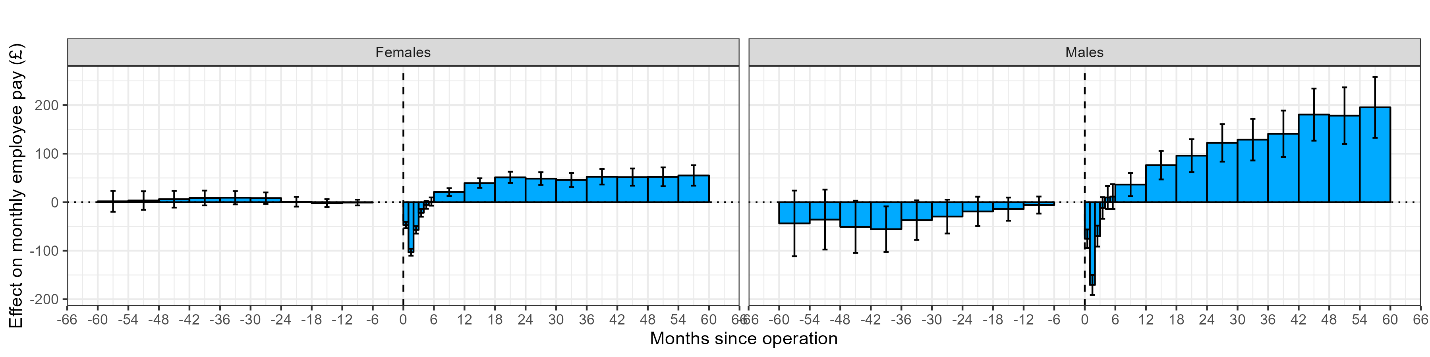

**Supplementary Figure 9:** Effect of bariatric surgery on monthly employee pay among those in work before and after surgery, by sex

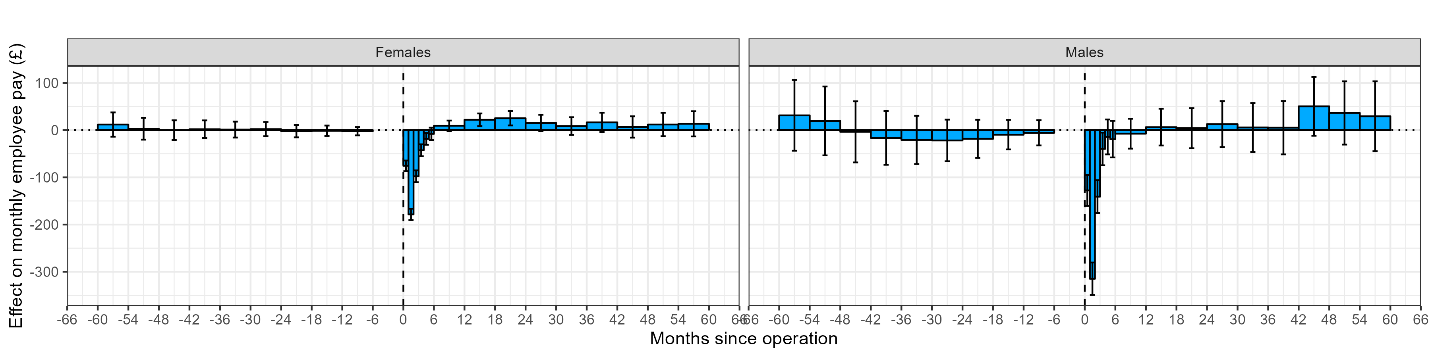

**Supplementary Figure 10:** Effect of bariatric surgery on probability of being a paid employee before and after surgery, by sex

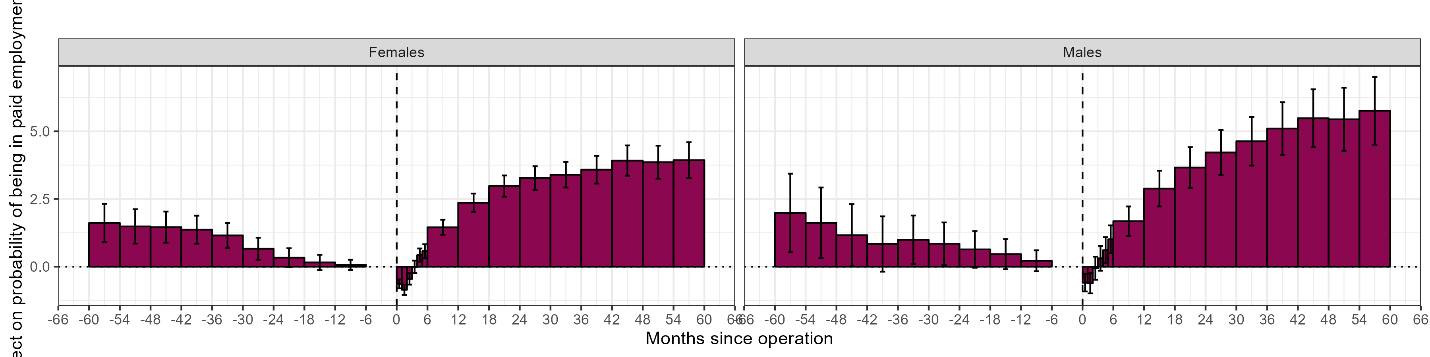

**Supplementary Figure 11:** Effect of bariatric surgery on monthly employee pay (overall) before and after surgery, across ethnic groups
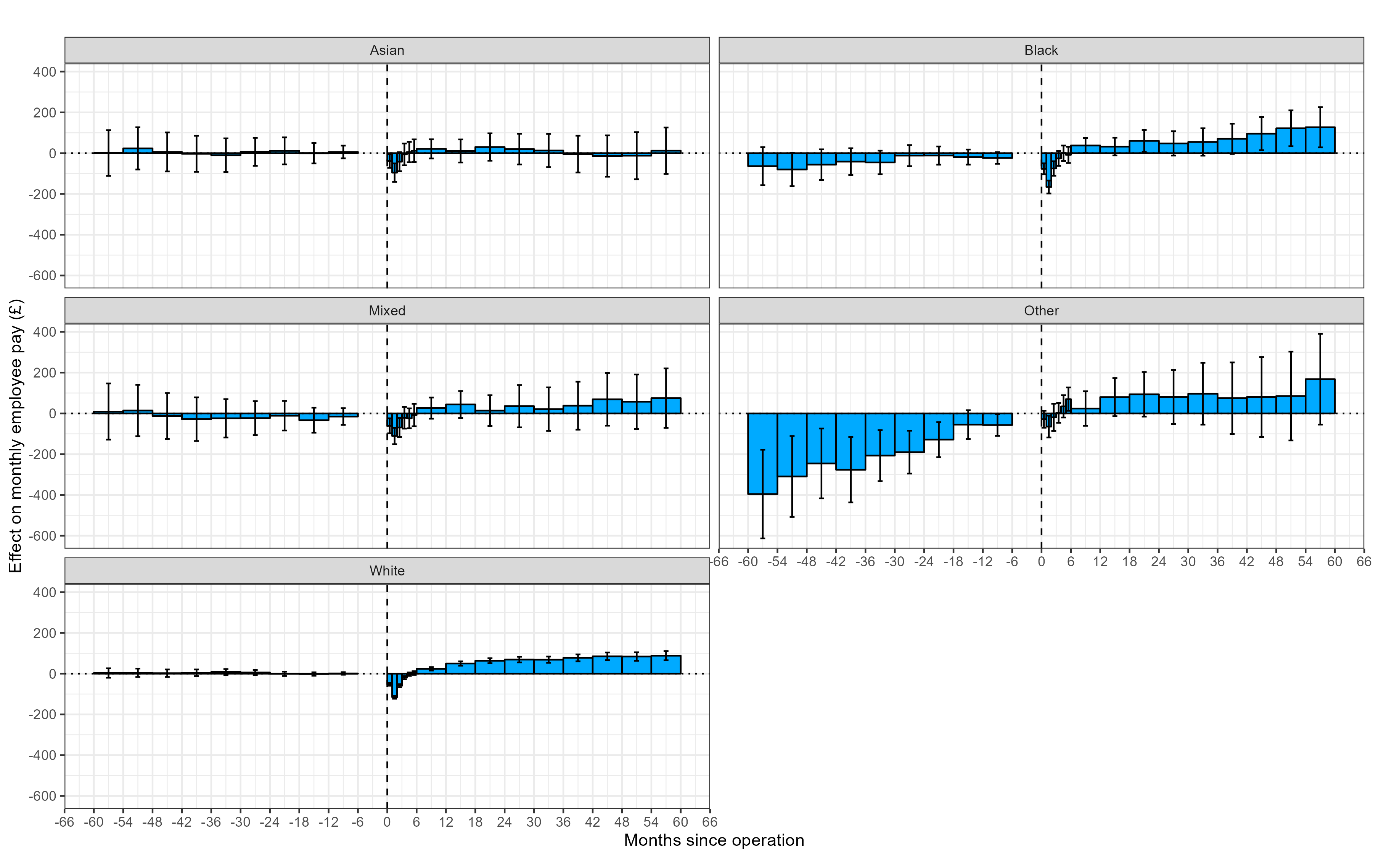

**Supplementary Figure 12:** Effect of bariatric surgery on monthly employee pay among those in work before and after surgery, across ethnic groups

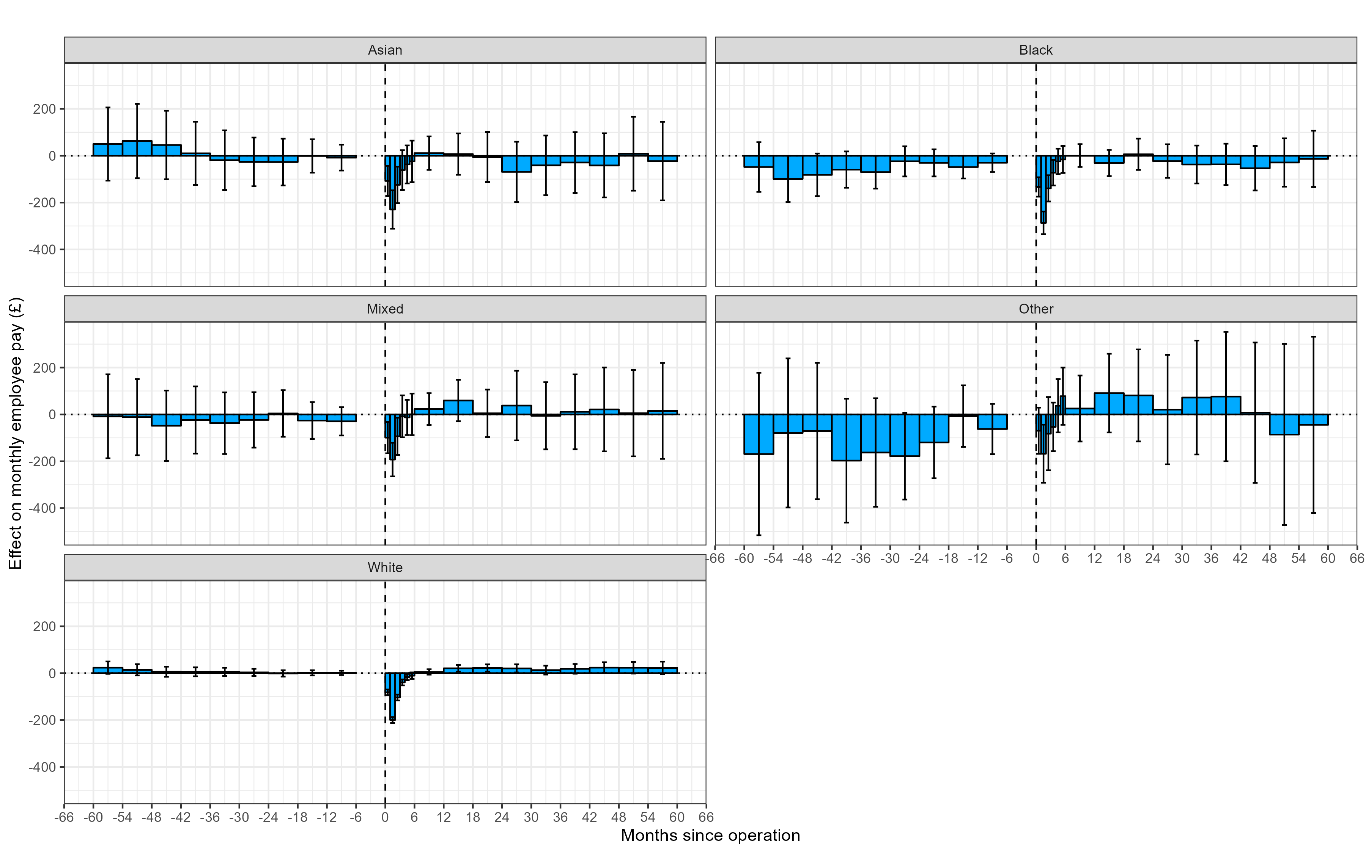

**Supplementary Figure 13:** Effect of bariatric surgery on probability of being a paid employee before and after surgery, across ethnic groups

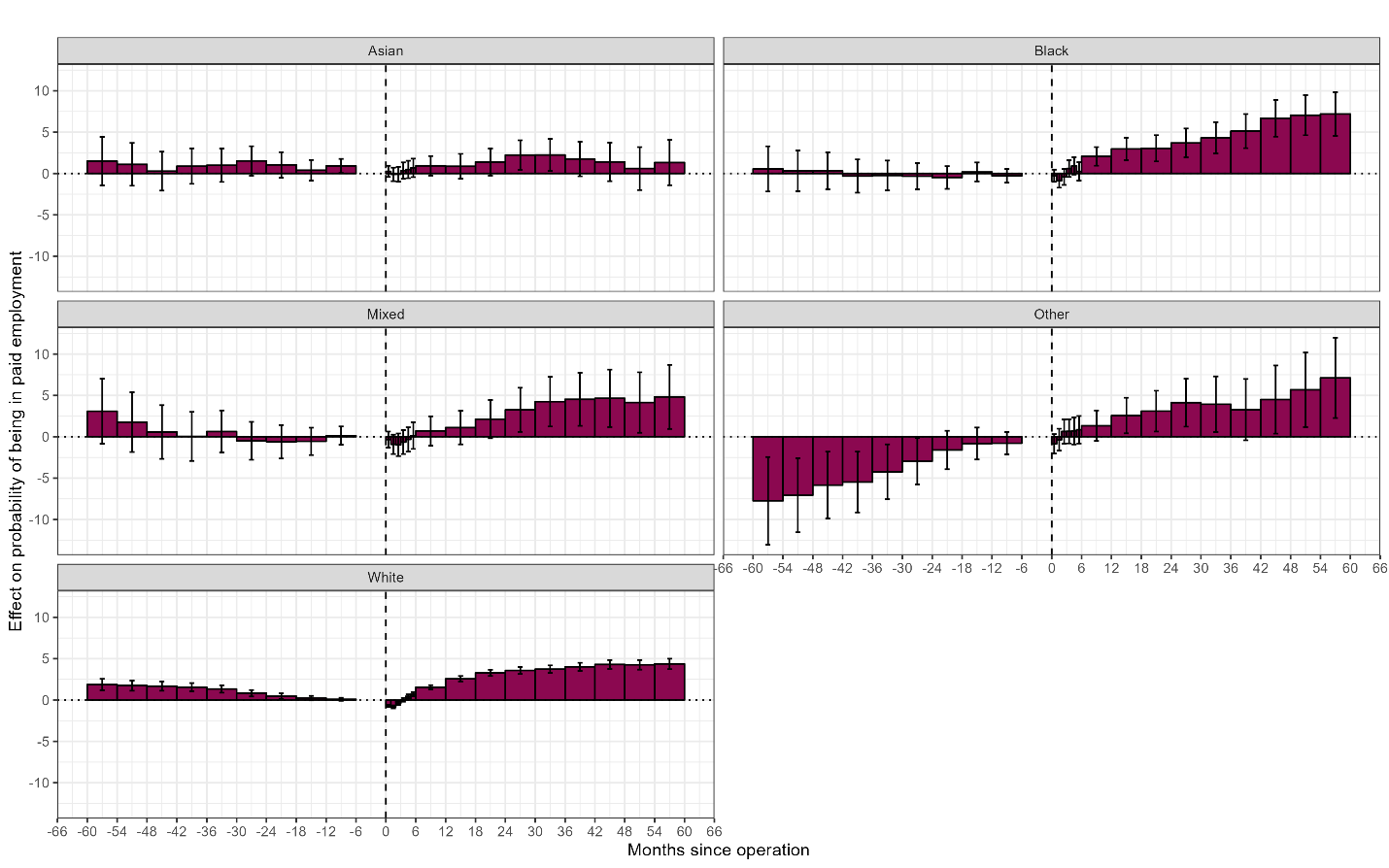

**Supplementary Figure 14:** Effect of bariatric surgery on monthly employee pay (overall) before and after surgery, across Index of Multiple Deprivation quintiles

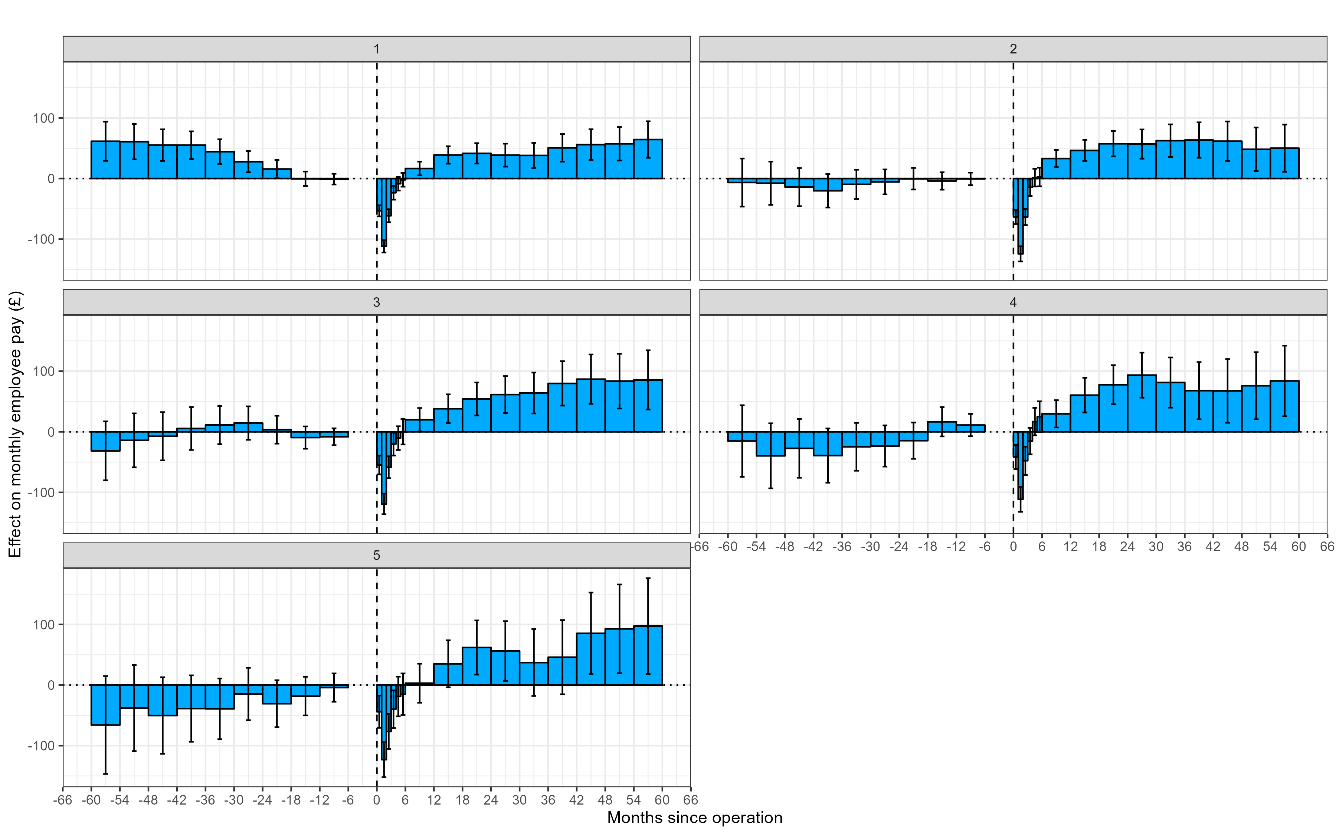

**Supplementary Figure 15:** Effect of bariatric surgery on monthly employee pay among those in work before and after surgery, across Index of Multiple Deprivation quintiles

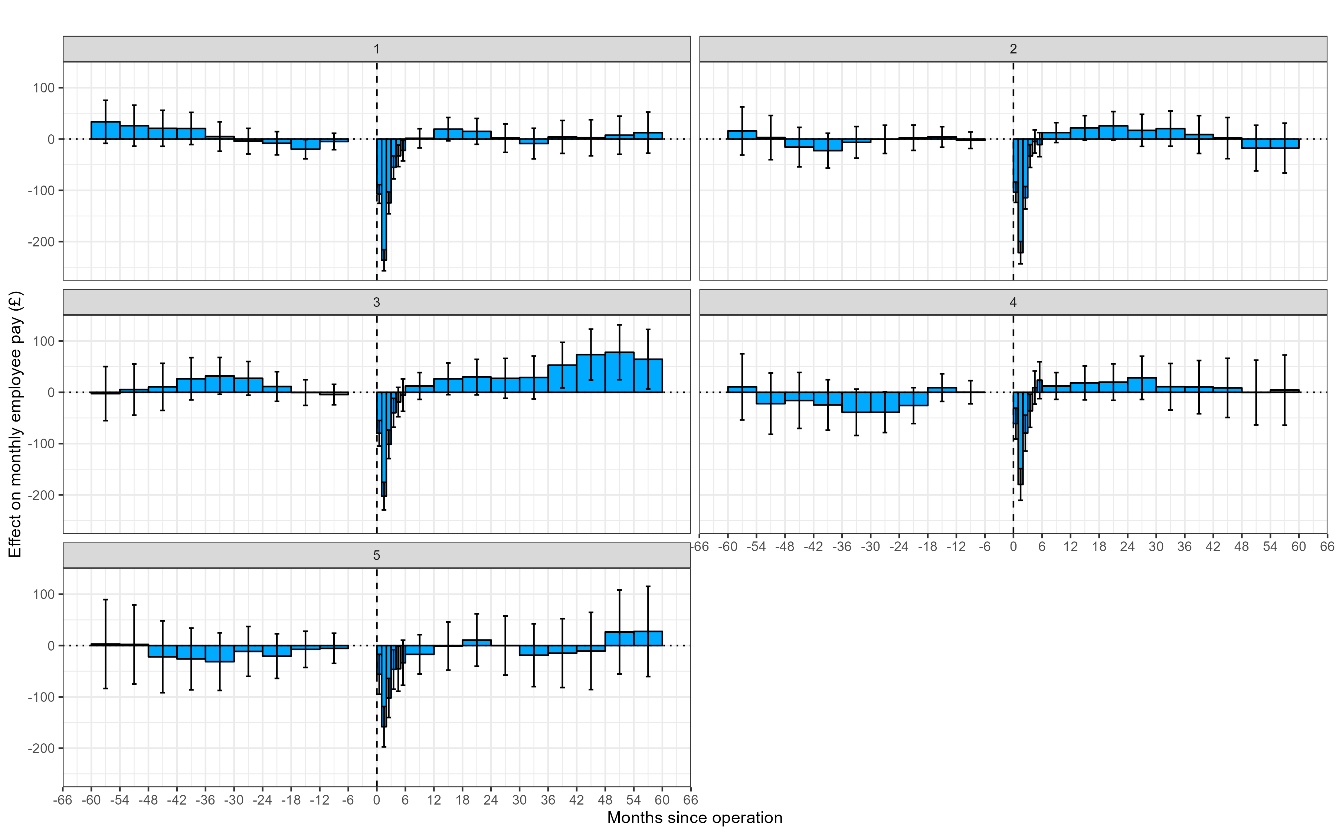

**Supplementary Figure 16:** Effect of bariatric surgery on probability of being a paid employee before and after surgery, across Index of Multiple Deprivation quintiles

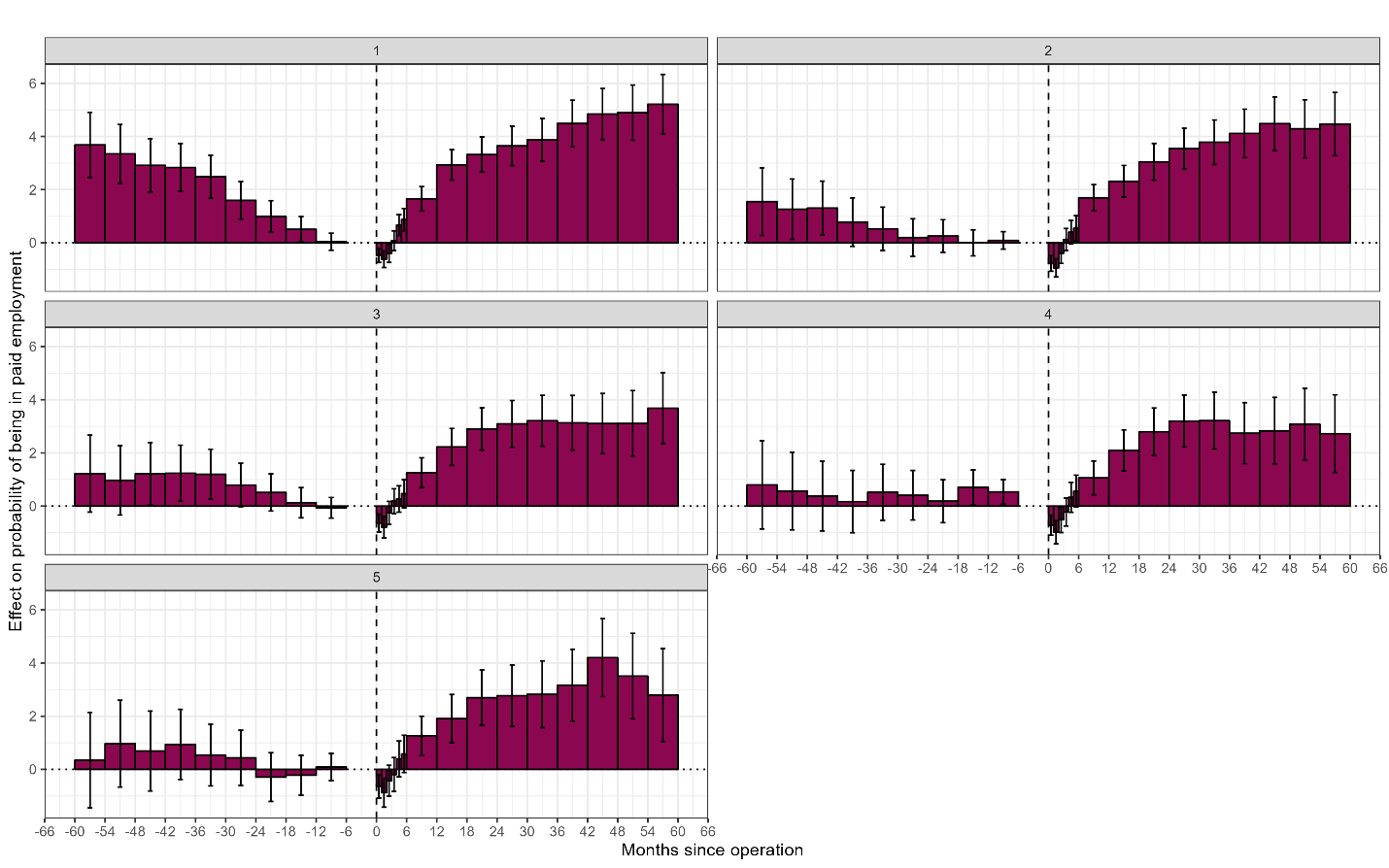

**Supplementary Figure 17:** Effect of bariatric surgery on monthly employee pay (overall) before and after surgery, across regions

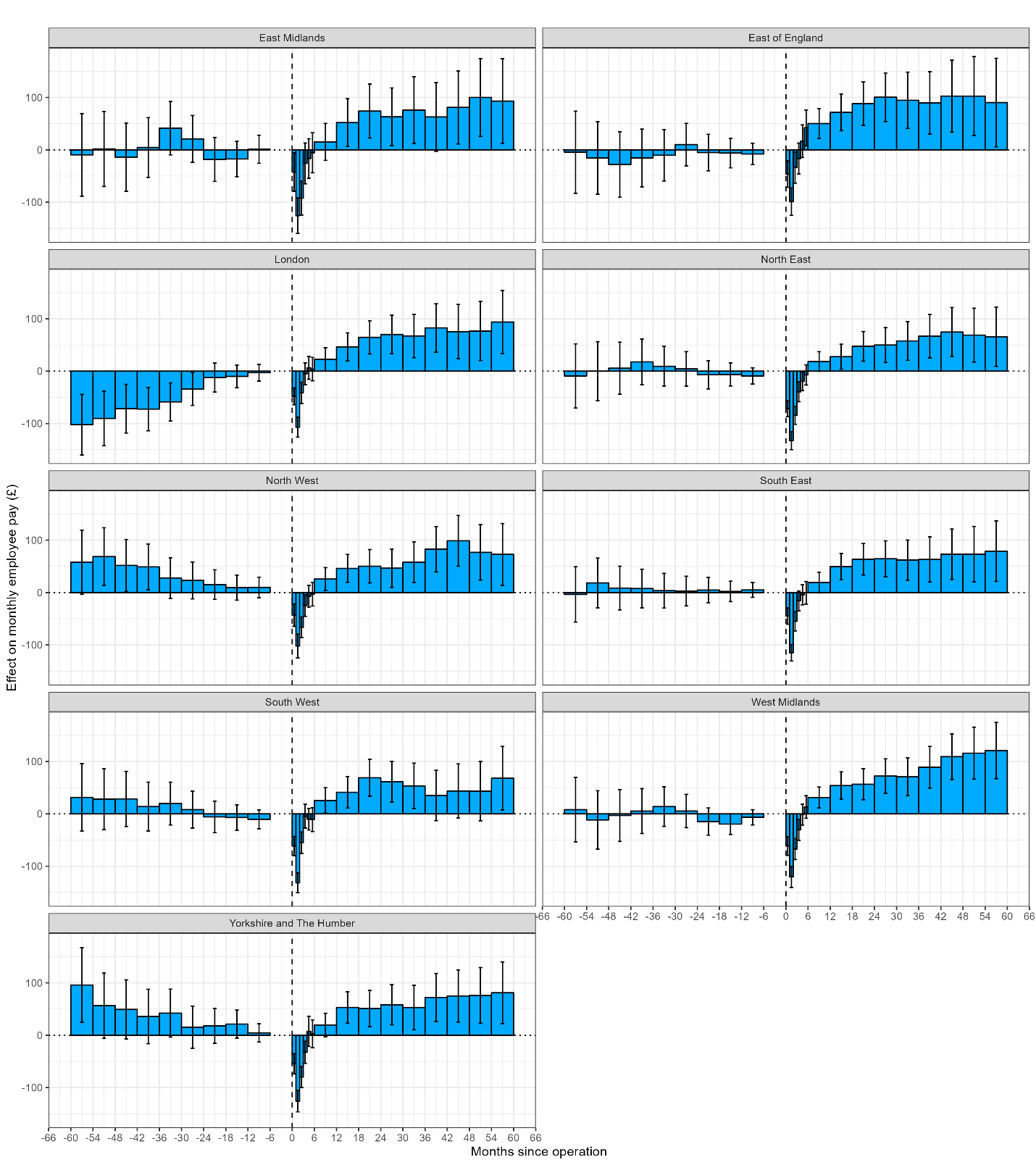

**Supplementary Figure 18:** Effect of bariatric surgery on monthly employee pay among those in work before and after surgery, across regions

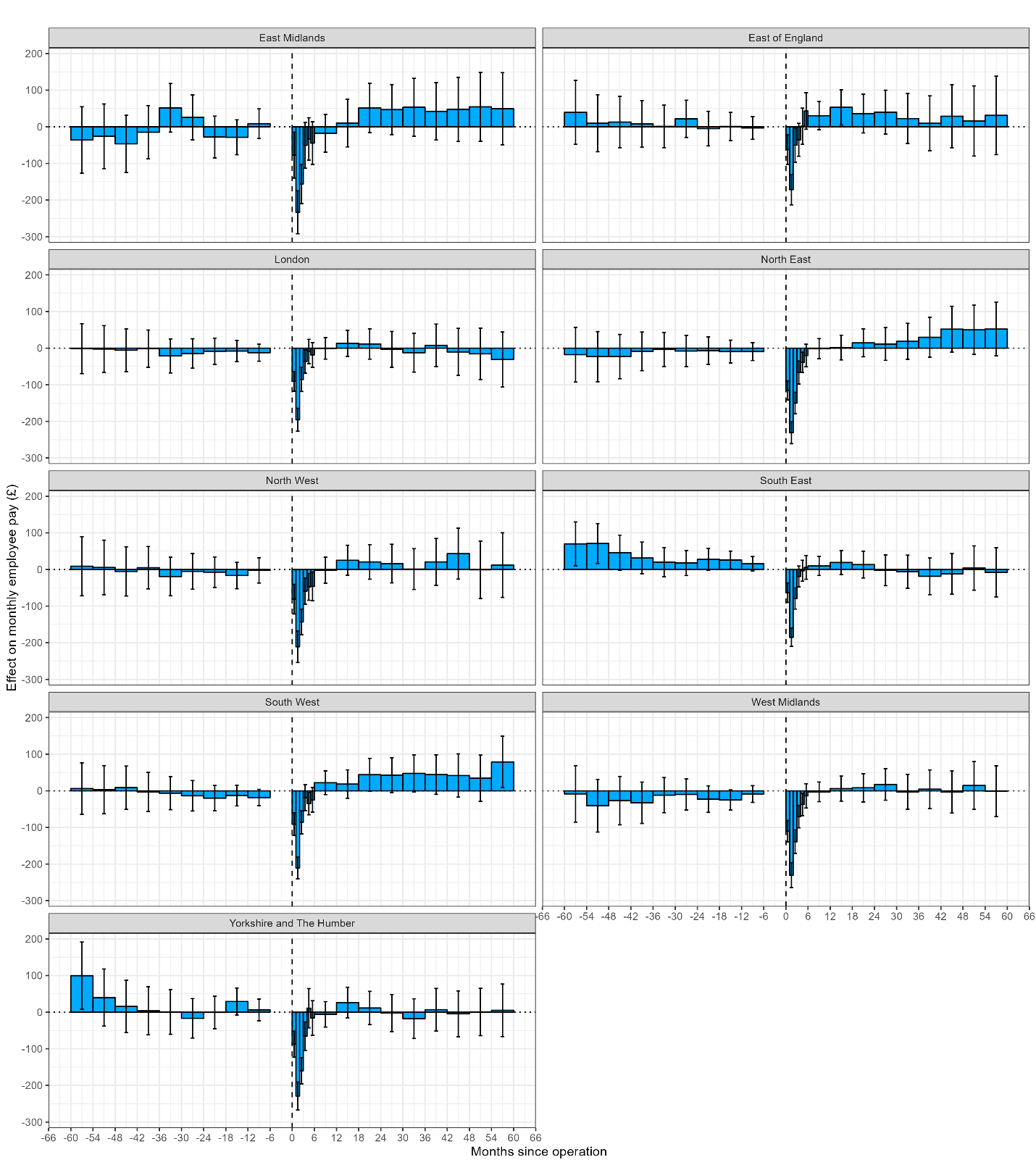

**Supplementary Figure 19:** Effect of bariatric surgery on probability of being a paid employee before and after surgery, across regions

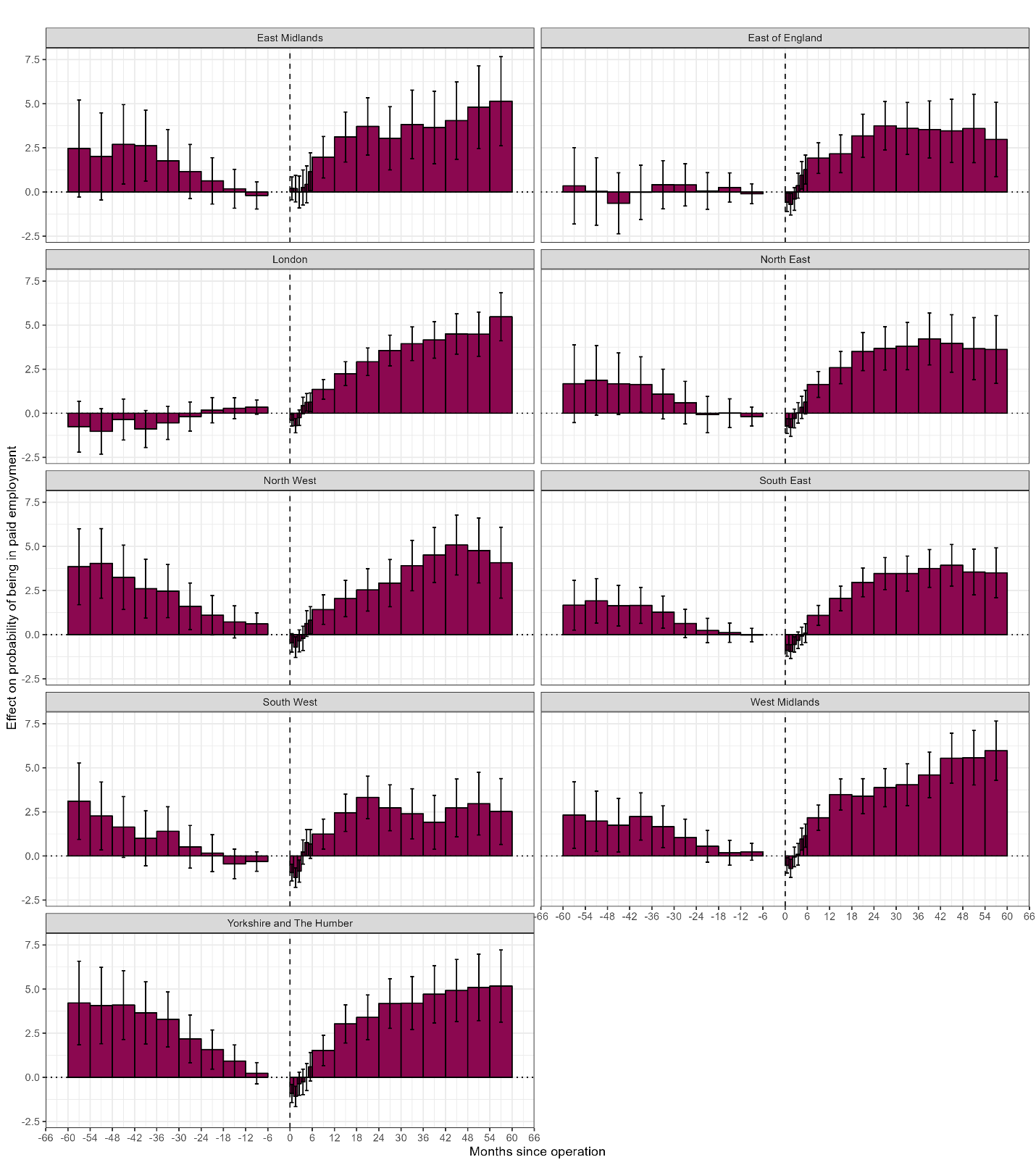

**Supplementary Figure 20:** Placebo test censoring at surgery

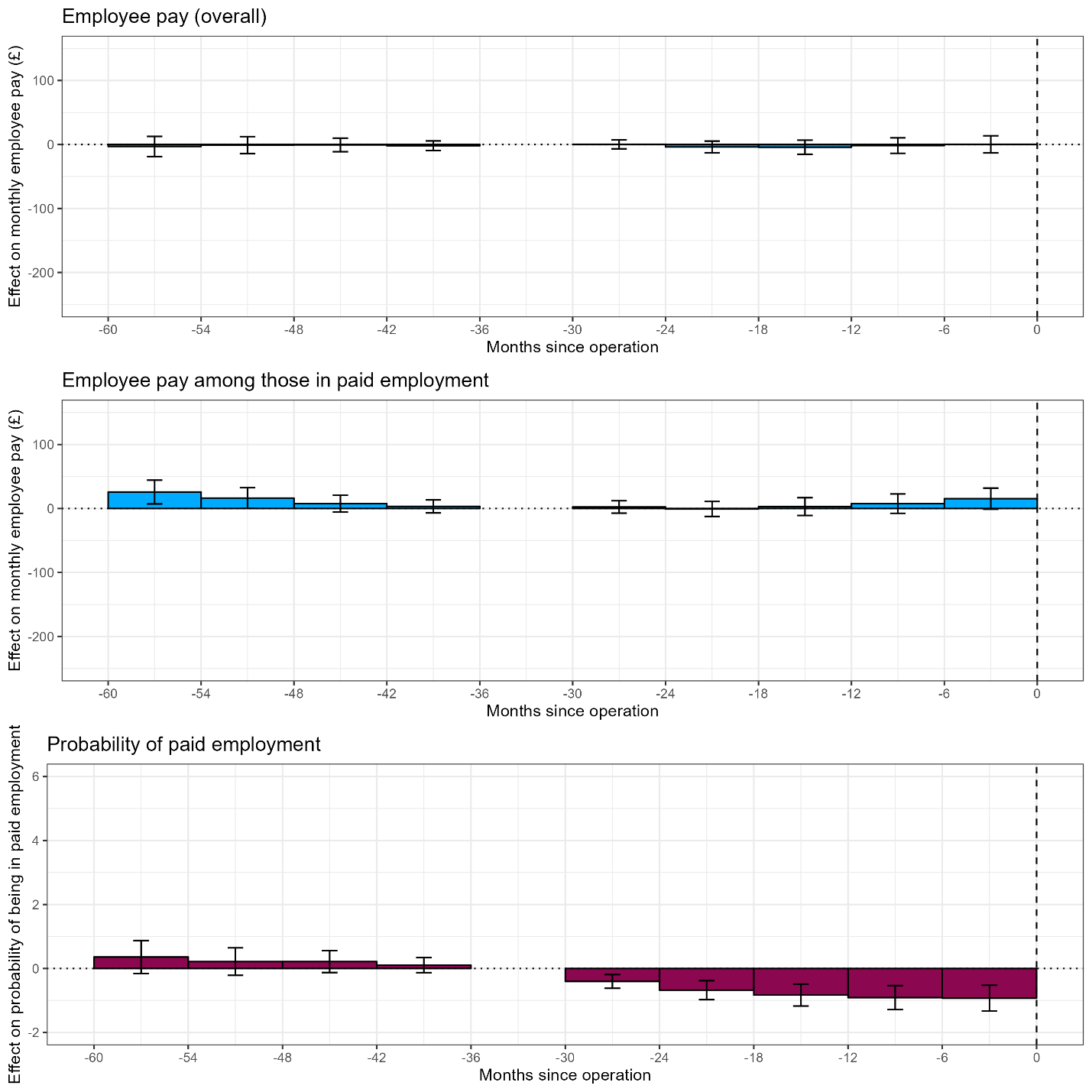

**Supplementary Figure 21:** Placebo test with unexposed only

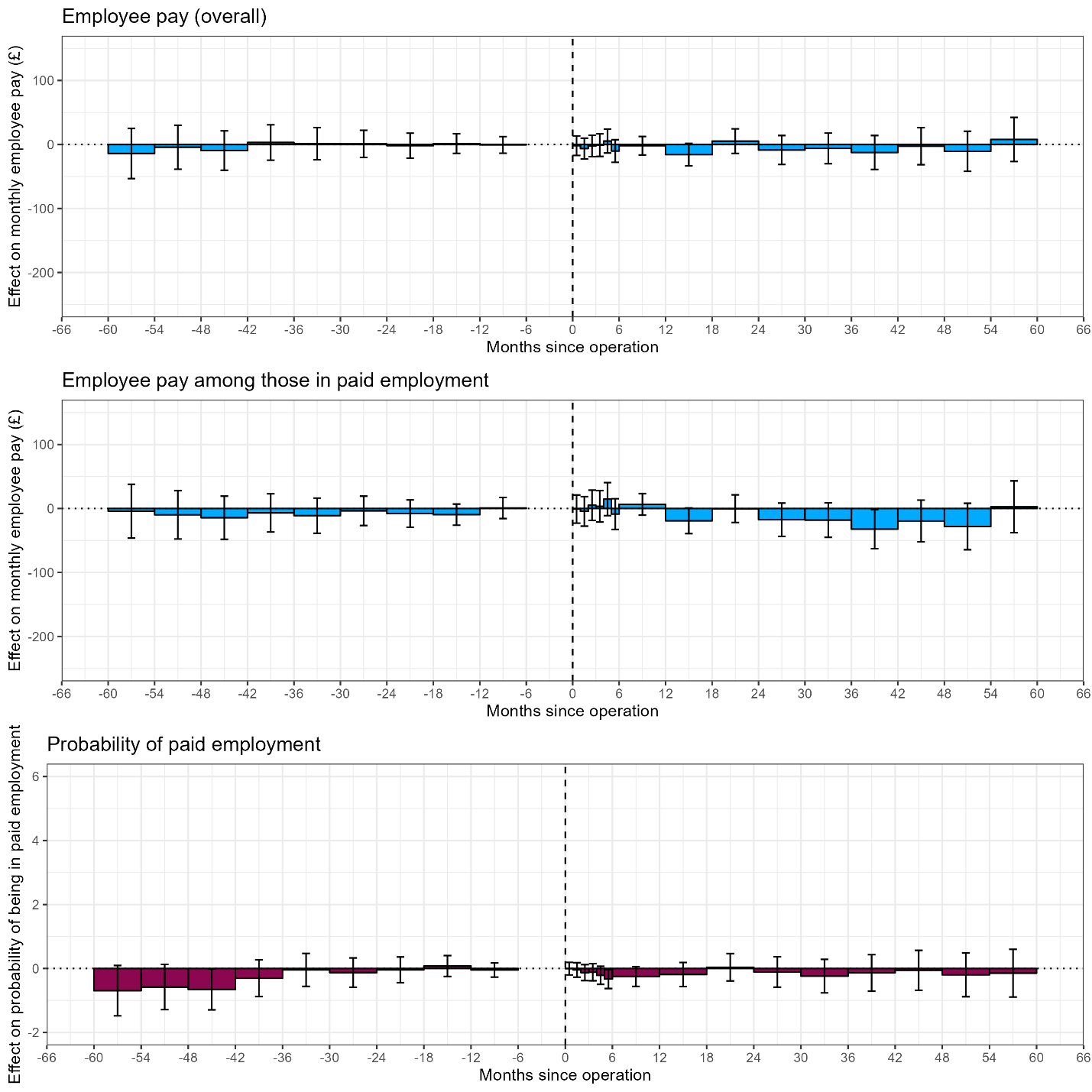
